# Mortality burden of outdoor occupational heat exposure in the United States

**DOI:** 10.64898/2026.01.29.26345131

**Authors:** Abas Shkembi, Leah H. Schinasi, Devon Payne-Sturges, Richard L. Neitzel

**Affiliations:** Department of Environmental Health Sciences, University of Michigan School of Public Health, Ann Arbor, MI, USA; Department of Environmental & Occupational Health, Dornsife School of Public Health, Drexel University, Philadelphia, Pennsylvania, USA; Urban Health Collaborative, Dornsife School of Public Health, Drexel University, Philadelphia, Pennsylvania, USA; Center of Occupational Health and Safety Engineering, University of Michigan, Ann Arbor, MI, USA

## Abstract

**Background:** Outdoor workers are particularly vulnerable to the adverse impacts of heat, but many studies focus on heat exposure in residential settings only. This leads to a limited understanding of the full mortality burden due to occupational heat exposures. Here, we aimed to improve estimates of the total, short-term mortality burden attributable to outdoor occupational heat exposure in the United States (US).

**Methods:** We developed a panel data set for 3,108 US counties during 2010-2019 by linking all-cause mortality among the working age population, derived from CDC WONDER, with the prevalence of workers exposed to outdoor occupational heat, which integrates data on wet bulb globe temperature, workplace activities, and employment counts. We developed a quasi-Poisson regression model adjusted for ambient temperature, total precipitation, and county and state-year fixed effects to estimate short-term excess deaths attributable to outdoor occupational heat exposure.

**Findings:** Nationwide, approximately 3.8% (95% CI: 2.5-5.8%) of all workers were annually exposed to dangerous wet-bulb globe temperatures. This outdoor occupational heat exposure resulted in approximately 9,800 (3,100-17,000) annual excess deaths in the working age population. An estimated 62% of excess deaths occurred in the most socially vulnerable counties despite accounting for 25% of workers.

**Interpretation:** The mortality burden of occupational heat exposure is likely far larger than 39 officially reported annual deaths that the Bureau of Labor Statistics reports for this time period. The workplace should be an explicit focus of heat policies, advocacy, and adaptation measures.

**Funding:** US Centers for Disease Control and Prevention/National Institute for Occupational Safety and Health.

## Introduction

Despite the well-established relationship between ambient heat exposure and mortality risk,^1^ the total mortality burden that heat represents to workers remains poorly understood. Most heat-mortality studies focus on vulnerable age demographics, such the elderly and children, due to their increased susceptibility to effects of heat exposure.^2^ While the working age population (defined as 15-64 years old in this study) is generally less biologically sensitive to the effects of heat because of their age and better underlying health, the intense heat exposures that some workers experience, sometimes while wearing bulky or heat-trapping clothing and engaging in high metabolic output activities, makes a subset particularly vulnerable to heat stress (i.e., effects of increased heat storage within the body).^3^

According to the U.S. Bureau of Labor Statistics, a third of all workers in the US regularly work outdoors,^4^ placing roughly 45 million workers at risk of heat-related mortality due to prolonged heat exposure (>8 hours), intense physical exertion, and limited options to avoid and control exposure (e.g., access to cooling). This has spurred efforts to protect workers from heat stress in the workplace, as evidenced by efforts to promulgate a nationwide occupational heat standard in the US.^5^ However, the 30-60 annual deaths officially attributed to occupational heat exposure^6^ represent a miniscule fraction of the >150,000 total annual deaths that are estimated to be attributable to high temperatures in the US.^7–9^ The relatively small fraction of total heat deaths that appear to be attributed to occupational heat has led most heat adaptation policies focus primarily on residential exposures among older adults.^10^

It has been hypothesized, however, that these yearly 30-60 work-related deaths are a substantial undercount of the total deaths due to occupational heat exposure (**Figure 1**).^11^ Heat stress occurs when the human body is not able to effectively maintain a healthy and safe internal temperature.^12^ Recorded heat-related deaths likely have obvious signs of having occurred as a result of core body overheating; for example, overheating often occurs in unacclimatized workers who are new to the job. Yet occupational heat can lead to death through a variety of pathways, such as ones that are kidney-, respiratory-, cardiovascular, injury- or psychiatric-related, and the deaths may present as such. This can lead physicians, medical examiners, and coroners to not recognize occupational heat as a cause of death.^11,13^ Additionally, many workers who engage in jobs that involve intense heat exposures are socially vulnerable to heat; this positionality and the additional stressors that they experience both inside and outside of their work environment can worsen the impacts of heat.^14^ It is thus critical to identify enhanced occupational heat-vulnerability in socially vulnerable populations.

**Figure 1.**
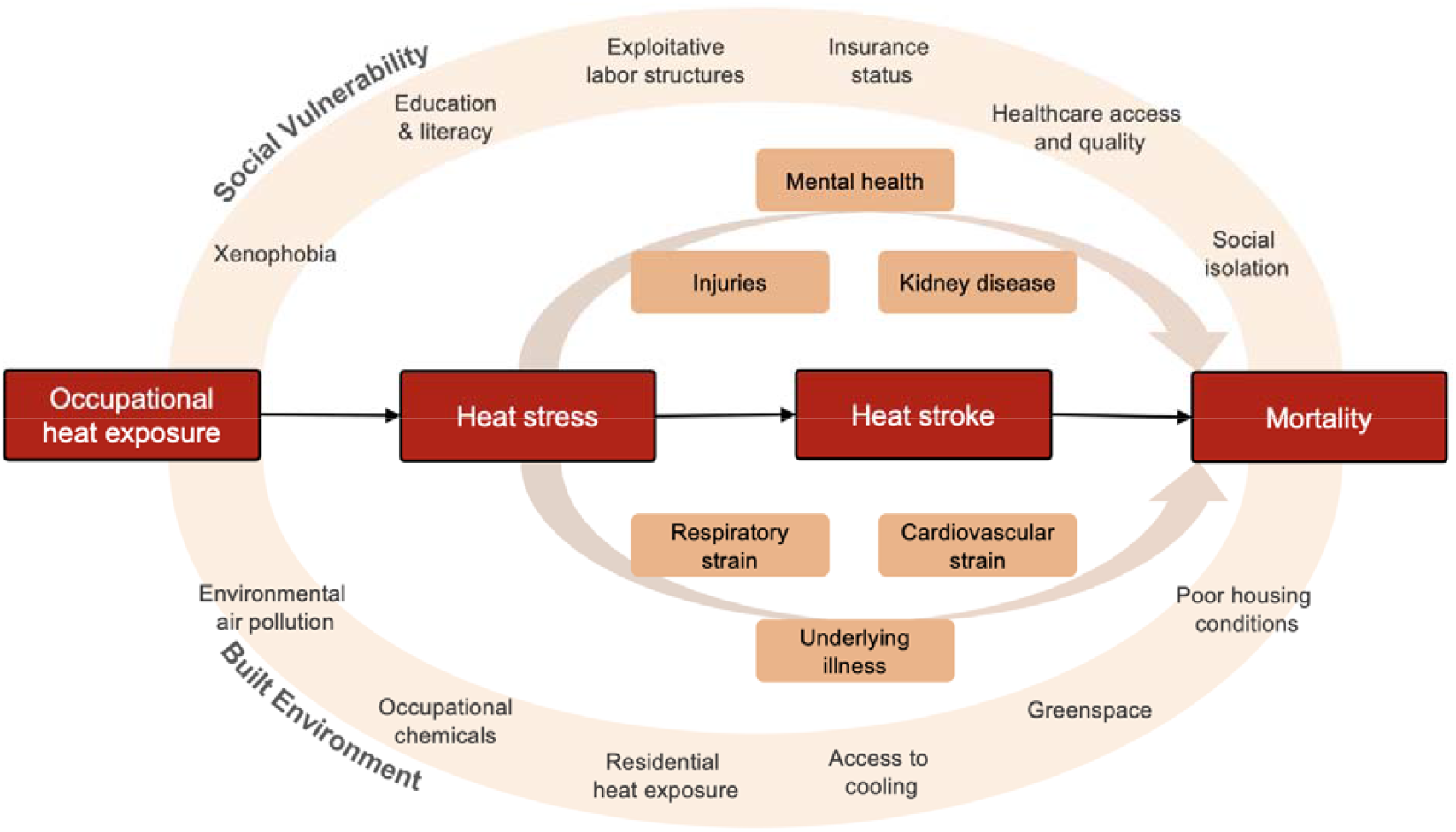
Conceptual diagram demonstrating the acute impacts of occupational heat exposure on mortality.

Here, we studied the short-term burden of occupational heat exposure on all-cause mortality among the working age population across 3,108 counties in the contiguous US (CONUS) during 2010-2019. We empirically estimated changes in the yearly county mortality rates due to occupational heat exposure, after accounting for non-work extreme heat, precipitation, and other unmeasured spatial and temporal confounders. By modeling all-cause deaths, rather than deaths classified as due to heat exposure or reported by worker’s compensation/mandated occupational reported data, we can comprehensively capture all deaths related to occupational heat exposure without requiring that every pathway is explicitly modeled, as work-related death estimates likely substantially undercount the total burden of occupational exposures.^15^

## Methods

### Occupational heat exposure

We estimated the percentage of workers likely exposed to hazardous levels of outdoor heat at work for every CONUS county annually during 2010-2019. Our exposure estimation approach was informed by previously published work.^16^ For this reason, we leave detailed methodology in Section 1 (pg. 9-14) of the Supplemental Materials and provide a brief overview of our approach below.

Conceptually, exposure was defined according to the American Conference of Governmental Industrial Hygienists (ACGIH) screening criteria for unacclimatized workers.^17^ Exposure reflects heat levels high enough to lead to some level of heat stress, but not always enough to lead to heat stroke, which is useful for capturing all pathways of heat mortality. Specifically, for each day during 2010-2019, we linked daily mean wet-bulb globe temperature level from Spangler et al. (2022)^18^ with detailed employment counts for each county from the US Bureau of Labor Statistics Occupational Employment and Wage Statistics,^19^ and augmented the data with factors related to heat stress vulnerability (working outdoors, metabolic rate, and allocation of work vs rest periods) from the US Department of Labor Occupational Information Network (version 24.1)^20^ to simulate the likely daily incidence of occupational heat exposure using a direct estimation approach (i.e., multiplying the daily probability of exposure with the total number of workers in a county).

This simulation yields the fraction of all workers exposed to occupational heat on day *d* within county *c* on year *y* during 2010-2019 (*F*_*dcy*_) as a function of working outdoors, ambient wet-bulb globe temperatures, metabolic rate, work/rest allocation, and occupational employment counts. We then applied this daily fraction estimate to estimate yearly prevalence while considering that the same workers could be exposed to heat on multiple days. To do so, we adapted and applied a previously developed method^21^ to estimate the overall exposure prevalence (*P*_*cy*_) for county *c* in year *y* when the variation in between-day exposure incidence is dependent

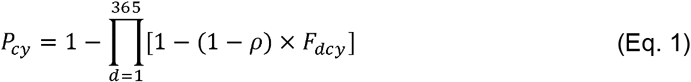

where *ρ* reflects the correlation in the number of workers exposed between each day. The true between-day exposure correlation is unknown, so we conservatively specify a time-invariant *ρ* = 0.99 to reflect that the workers exposed to heat each day are highly similar, but not always the same (i.e, *ρ* = 1).

### Mortality outcome

We obtained county-level, yearly underlying cause of death data from the US National Center for Health Statistics from 2010-2019 from the Centers for Disease Control and Prevention (CDC) WONDER platform.^22^ Ideally, we would only assess deaths where occupational heat was an underlying, contributing factor of death. However, since it is impossible for physicians, medical examiners, and coroners to always identify such cases, we assessed all-cause mortality, as is common in the heat-mortality literature.^23^ We did not assess specific causes of death since our goal was to estimate the total mortality burden of occupational heat exposure, and because heat can lead to mortality through a variety of mechanisms.^1^ While older adults 65+ years old often work, we assessed mortality among the working age population of 15-64 years only to minimize bias from unknown, non-work-related factors from older age and since older adults are underrepresented in jobs at greatest risk of heat exposure.^24^

### Covariates

To assess if rates of occupational heat deaths vary according to social vulnerability, we used county-level data from the CDC Social Vulnerability Index (SVI) for 2016.^25^ The 2016 date served as a near-midpoint of the study period. The SVI incorporates data on socioeconomic status, household characteristics, racial and ethnic minority status, housing type, and transportation to calculate a percentile from 0-1, where 1 indicates the county with the highest relative vulnerability in the country.

To account for confounding from non work-related heat exposure, we additionally estimated the annual number extreme heat days. For each county-year, extreme heat was defined as days with daily wet-bulb globe temperature higher than the county’s 90th percentile warm season temperature (May to September, 2010 to 2019) and the days were summed.^18^ The 90^th^ percentile threshold was arbitrarily selected to reflect exposure to extreme heat days within a county. Precipitation may also confound the association between heat and mortality. Thus, we also included as a covariate yearly, mean precipitation (in inches) for each county in the US between 2010-2019 from the US National Oceanic and Atmospheric Administration (NOAA) National Centers for Environmental Information.^26^

### Modeling occupational heat and mortality relationship

Analyses were conducted in R v4.5.1. We used a quasi-Poisson, panel fixed effects regression models to assess the short-term effect of occupational heat exposure prevalence on county-level all-cause mortality rates. The quasi-Poisson model specification accounts for overdispersion of mortality counts. Panel fixed effects model have been widely used in econometric studies^27^ and have been increasingly applied to environmental epidemiology studies.^28,29^ We model the percent change in all-cause mortality rates for county *c* within state *s* and year *y* for every unit change in the percent of workers exposed to occupational heat (%*Exp*_*csy*_), using the following model specification:

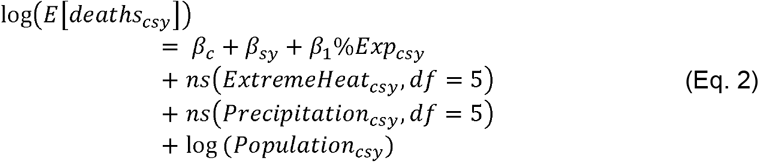

where *deaths*_*csy*_ is the number of all-cause deaths among 15-64 year olds; *β*_*c*_ is the county-specific intercept that accounts for any county-specific, time-invariant factors that could be associated with both occupational heat exposure and mortality (e.g., industrial composition); *β*_*sy*_ is the state-year specific intercept used to control for factors that vary across state and time (e.g., policies); *β*_1_ is the main coefficient of interest for the percentage of workers likely exposed to occupational heat at work; *ns* (*Extreme Heat*_*csy*_, *df* = 5) and *ns* (*Precipitation*_*csy*_, *df* = 5) are natural spline terms with an arbitrary 5 degrees of freedom to account for the time-varying effect of non-occupational heat exposure and precipitation on mortality; log (*Population*_*csy*_) is the natural logarithm of total population of 15-64 year olds, included as a population offset. We ran an additional model stratifying occupational heat exposure by 2016 SVI quartiles (4th quartile reflecting the most vulnerable counties) to explore potential heterogeneous effects due to social vulnerability.

Our modelling approach here leverages within-county, year-to-year variation in occupational heat (**Supplemental Figure S1**) to estimate the short-term effect of outdoor workplace heat exposure on all-cause mortality. This approach minimizes confounding bias due to time-invariant factors that are unlikely to vary substantially within counties over this time period (e.g., industrial composition, rurality, general socioeconomic conditions, age composition). Additionally, our approach accounts for factors that differ across states in a given year (e.g., occupational heat stress regulations) and for potential time-varying confounding bias resulting from non-occupational heat or precipitation exposure. The within-county variation of the prevalence of occupational heat exposure is also driven by stochastic atmospheric changes year-to-year, minimizing bias due to unmeasured time-varying confounders.

### Estimating mortality burden

We quantified the number of short-term excess deaths attributable to outdoor occupational heat exposure (Δ*deaths*_*cy*_) for county *c* in year *y* using the following, previously established formula:^30^

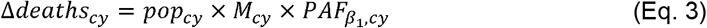

where *pop*_*cy*_ denotes the size of the working age population (15-64 years old), *M*_*cy*_ is the all-cause mortality rate for the population, and 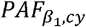 is the population attributable fraction associated with the change in percentage of the population exposed to occupational heat, defined as

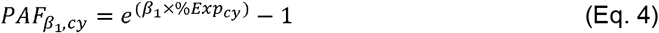

such that %*Exp*_*cy*_ reflects the counterfactual where no workers were exposed to occupational heat for a given county-year, independent of ambient, extreme heat exposure. We assessed the composition of the estimated excess heat deaths by social vulnerability (four quartiles), US state, and US region (Northeast, Midwest, South, and West, as defined by the US Census Bureau).

### Sensitivity analyses and robustness checks

Our analysis relies on the assumption that the observed occupational heat-mortality relationships estimated from our regression model can be interpreted as plausibly causal. We conducted several additional analyses and robustness checks to test the sensitivity of our results to assumptions related to causal inference. Specifically, we probed the quality of our data, whether we correctly specified the regression model, and whether we likely violate assumptions of exchangeability and consistency. Specific details of the analytical approaches are presented in the Supplemental Materials (Section 2; pg. 14-21).

## Results

### Summary

There were approximately 669,000 annual all-cause deaths among the working age population during 2010-2019. During this time period, on average, an estimated 5.7 million workers nationwide (95% CI: 3.8-8.5 million) were exposed to outdoor occupational heat each year, translating to an annual average of 3.8% (95% CI: 2.5-5.8%) of workers. Outdoor occupational heat exposure did not vary substantially year-to-year nationwide (**Table S1**), but did vary by county (**Figure 2a; Figure S1**). Roughly a sixth of counties, primarily in the Southern US, had >10% of workers exposed to heat, and 17 counties in Florida, Texas, and Louisiana had >20% of workers exposed. More socially vulnerable counties were more exposed to occupational heat than less socially vulnerable counties (**Figure 2b**). For example, counties in the fourth SVI quartile had a median of 8.6% (IQR: 3.2-11.7%) of workers annually exposed, while the counties among the first quartile had a median of 1.1% (IQR: 0.2-2.4%).

**Figure 2.**
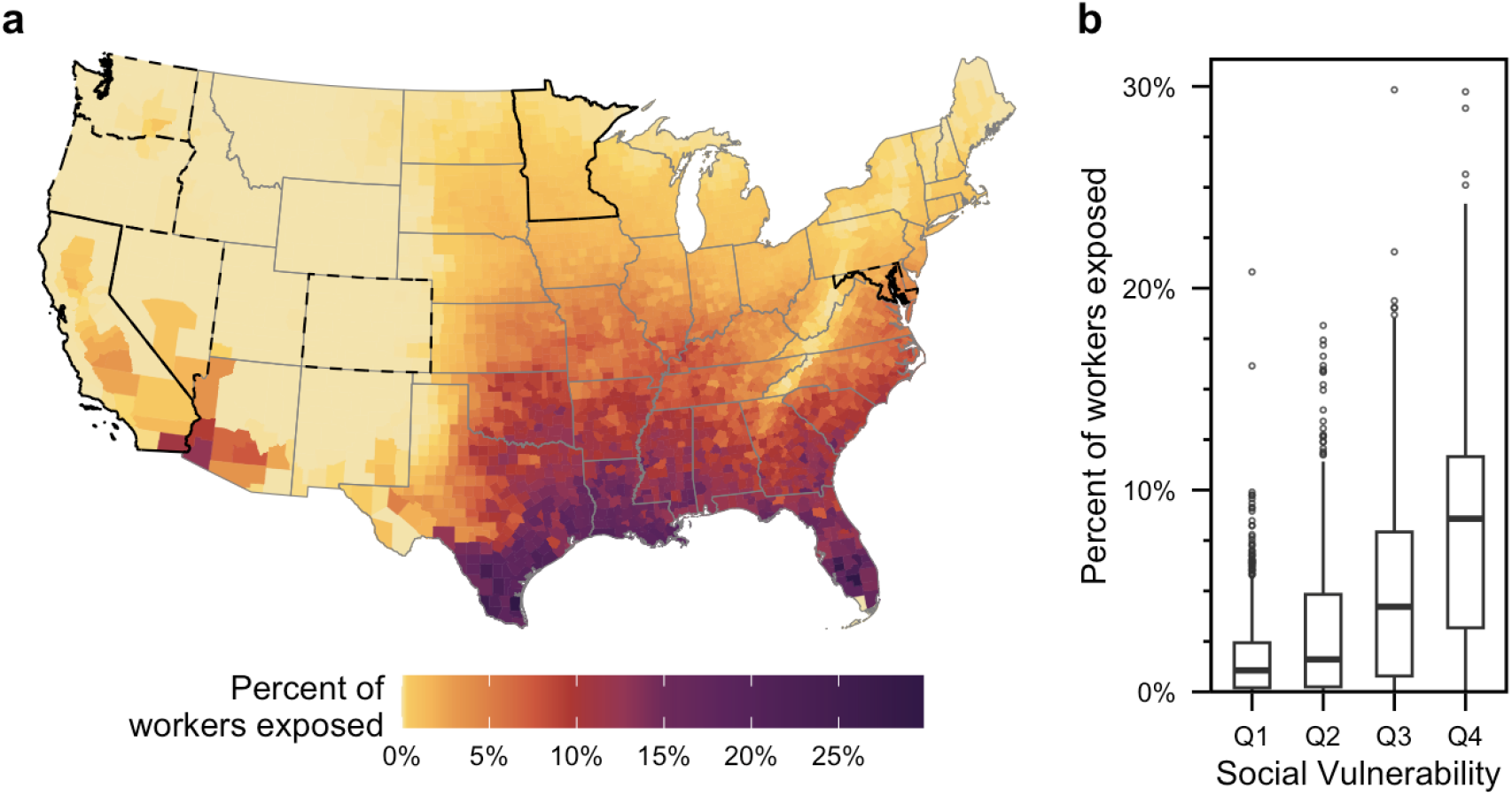
Spatial variability in occupational heat exposure and social vulnerability. **(a)** Estimated percentage of workers exposed to occupational heat during 2010-2019 in the contiguous United States by county. States with solid black border had implemented an occupational heat stress standard by 2019 (CA and MN); those with dashed black border had implemented a standard by 2025 (WA, OR, NV, CO, MD). **(b)** Boxplot showing distribution of county-level percentage of workers exposed to occupational heat (averaged across 2010-2019), stratified by categories of county-level social vulnerability (quantified using the CDC’s social vulnerability index, categorized in quartiles where highest quartile indicates highest social vulnerability in 2016).

### Relationship between occupational heat and mortality

In non-stratified analyses, we observed 3.3% (95% CI: 1.0-5.6%) higher county-level all-cause mortality rate for every 10% increase in percentages of workers exposed. This equates to around 1.7 deaths per 1,000 workers exposed. The magnitude of this association was stronger in the most socially vulnerable counties (**Figure 3a**), where the most socially vulnerable counties had a 3.3% (95% CI: -1.9-8.7%) higher mortality risk than the least socially vulnerable counties per 10% increase in workers exposed (**Table S2**).

**Figure 3.**
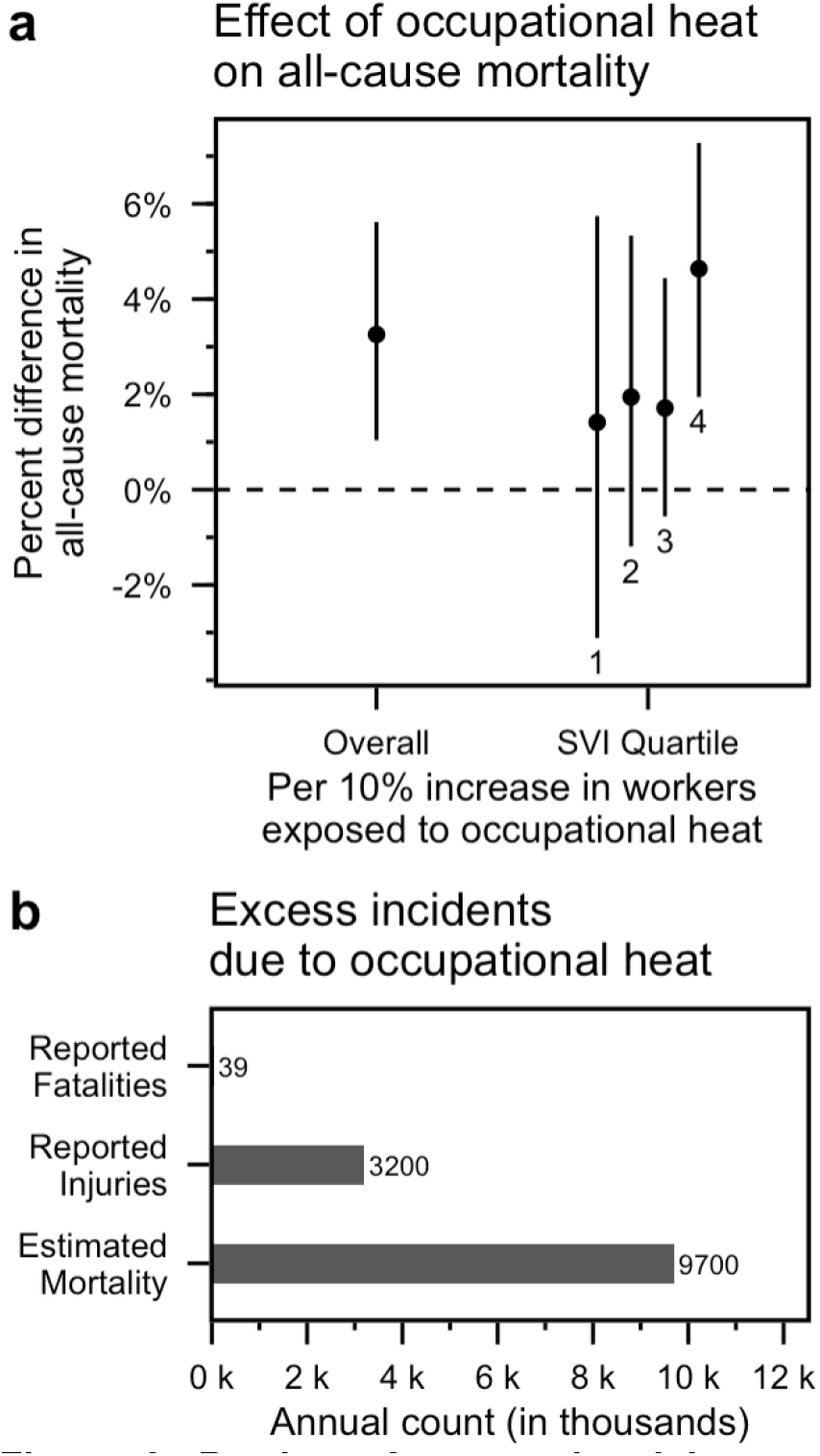
Burden of occupational heat exposure on all-cause mortality among the working age population. **(a)** Empirically estimated effects of the percentage of workers exposed to occupational heat (x-axis) on county-level, all-cause annual mortality rates among 15-64 year Americans during 2010-2019, across all US counties and stratified by 2016 social vulnerability index quartiles. A higher quartile reflects a higher degree of social vulnerability. **(b)** Estimated annual excess deaths due to occupational heat exposure from this study compared to officially reported heat-related injuries and fatalities according to the Bureau of Labor Statistics.

### Estimated excess deaths

We estimate that, across all counties, occupational heat exposure was associated with 9,800 annual excess deaths (95% CI: 3,100-17,000) during 2010-2019 (**Figure 3b**). Nationwide, occupational heat accounted for 1.5% of all deaths in the working age population during 2010-2019. We observed substantial heterogeneity in estimated excess deaths. Excess deaths due to occupational heat were highest in Louisiana (5.5%) and Mississippi (5.1%) (**Figure 4a; Figure S2**). States in the Southern US together accounted for 86.0% of excess deaths while representing 37.7% of all US workers. Exposures among workers living in the most socially vulnerable counties (fourth SVI quartile) accounted for the majority of excess deaths (62.0%), despite accounting for 24.5% of workers (**Figure 4b; Table S3**).

**Figure 4.**
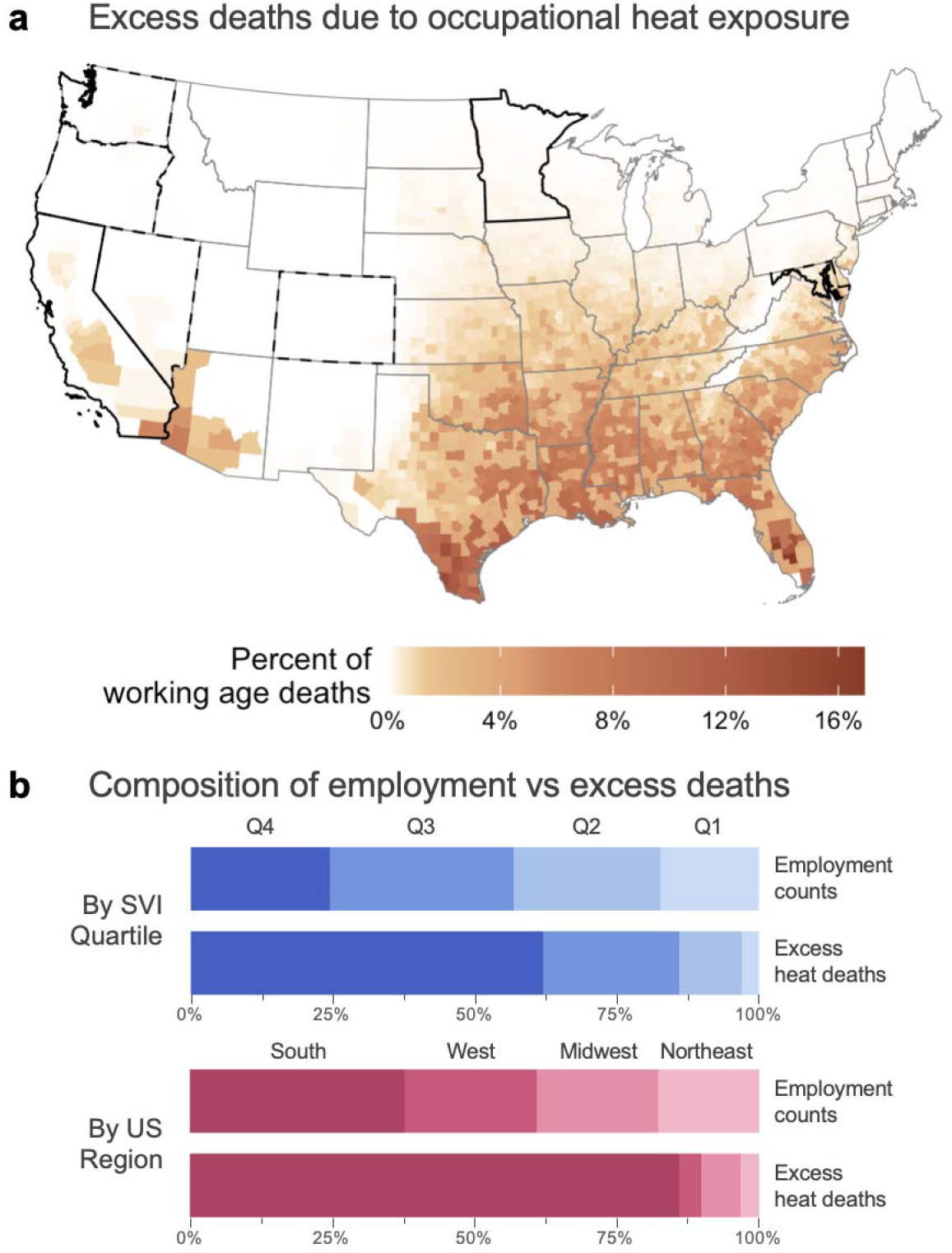
Geographic and sociodemographic patterns in the annual excess mortality due to occupational heat exposure. **(a)** Estimated, annual proportion of all-cause deaths among the working age population (15-64 years old) due to occupational heat exposure by county, averaged across 2010-2019. States with solid black border had implemented an occupational heat str ss standard by 2019 (CA and MN); those with dashed black border had implemented a standard by 2025 (WA, OR, NV, CO, MD). **(b)** Comparison between the composition of employment vs exc ss deaths by social vulnerability quartiles and US region. Top bars display the nationwide distribution of workers; bottom bars display the distribution of excess deaths due to occupational heat exposure. A higher SVI quartile indicates a higher degree of social vulnerability.

### Sensitivity analyses and robustness checks

Our results remain robust when probing their sensitivity to assumptions related to data quality, correct model specification, exchangeability, and consistency (Supplemental Materials, Section 2; pg. 14-21). Specifically, our specified quasi-Poisson, fixed effect regression model predicted mortality well (**Figure S8**), performed similarly to a negative binomial model (**Table S9**), and was not strongly influenced by how suppressed mortality counts were handled (**Table S8**). Three placebo tests did not produce spurious effects when using randomly reshuffled occupational exposure prevalence estimates across both county and year, within counties, or within year (**Figures S7, S9** and **S11**), providing additional evidence towards the validity of our exposure characterization, that our models adequately capture time-invariant confounders between counties, and that the models were not biased by spillover effects across counties. Controlling for additional, potential time-varying confounders (educational attainment, income, labor force participation, health insurance status, ambient air quality concentrations, and extreme cold exposure) did not change estimated effects, suggesting that estimates were not confounded by these variables (**Table S10**). An E-value sensitivity analysis further suggested that an unmeasured time-varying confounder, if one exists, would be unlikely to explain away the estimated occupational risk effects, conditional on the confounders already included and even the additional time-varying confounders tested in the sensitivity analysis (Supplemental Materials, pg. 19-20).

## Discussion

We found that occupational heat exposure is a meaningful driver of heat-related mortality in the working age population, particularly for socially vulnerable populations. We estimated that outdoor occupational heat exposure imposes an annual burden of 3,100-17,000 excess deaths, likely around 9,800 deaths. The most socially vulnerable areas in the US were both disproportionately exposed to occupational heat and impacted by excess occupational heat deaths.

A lack of previous estimates on the full, nationwide mortality burden of occupational heat limits our ability to assess the reliability of our estimates. The US Bureau of Labor Statistics reported around 30-60 annual occupational heat-related deaths each year during 2011-2019,^6^ while we find that the full mortality burden could by nearly 250 times higher. This difference is consistent with previous estimations of ambient heat mortality. The US CDC reports around 800 annual heat-deaths deaths among the total US population,^31^ yet previous studies have estimated that the total annual mortality burden of high temperatures could be around 150,000 deaths,^7–9^ nearly 185 times higher. This gap between officially reported statistics and our estimated mortality burden is plausible because reported occupational deaths likely reflect the most severe and acute impacts of heat, often where exposure and signs of heat strain are obvious, observed during work, and quickly lead to death.

The higher risk of heat-related mortality among adults ages 65 and older is well documented, and indeed, many studies of heat and mortality focus on this subpopulation.^2,32^ Our study suggests that occupational heat exposures in the working age population (15-64 years) may contribute more substantially to heat-related mortality than previously thought. Future work should consider incorporating occupational heat into studies of ambient heat to better capture mortality burden in the working age population, particularly in countries with less stringent regulations for occupational safety and health.

### Limitations

Many of the limitations of our study are likely to result in under-estimation of occupational heat exposure prevalence, thus potentially underestimating the full mortality burden. Our analysis examined occupational heat exposure among outdoor workers only and did not include additional, likely exposure among some indoor workers. Our approach to characterize the prevalence of workers exposed relied on making a conservative assumption that the workers exposed one day are very strongly correlated with the workers exposed the next day (*ρ* = 0.99 in Eq. 1). The true between-day correlation may be lower (e.g., *ρ* = 0.95), in which case our exposure prevalence is underestimated. Our exposure prevalence estimates were derived from data sources that include counted workers; since the most socially vulnerable workers are likely disproportionately burdened, not being able to incorporate uncounted workers (e.g., undocumented immigrants, incarcerated workers) likely further biases the total mortality burden low. Exposures were characterized according to a worker’s main job, missing additional exposures faced by temporary or gig workers with multiple jobs. Workers aged 65+ years can also be exposed to occupational heat, although we limited our characterization to the working age population (15-64 years old).

Our modeling approach also does not estimate longer-term health impacts of occupational heat that can take decades to manifest. Focusing on mortality only also excludes non-fatal health impacts from occupational heat, which are likely quite significant. Our work identified that occupational heat plausibly caused excess mortality in the working age population, but we cannot elucidate the particular, underlying mechanisms that result in excess mortality. While we found that more socially vulnerable areas are disproportionately burdened by excess occupational heat mortality, our data limits inference regarding whether certain sociodemographic groups (e.g., by race and ethnicity) experience health disparities as a consequence.

Our modeling approach can nonetheless result in biased estimation (either lower or higher) for individual counties. Our estimations are susceptible to the modifiable areal unit problem given that counties are arbitrary administrative boundaries. Heat is derived from average, ambient wet-bulb globe temperatures and does not capture hourly variation in heat. Our modeling does not account for individuals who migrate out of their county throughout the year due to environmental disasters, such as tropical cyclones.

### Public health implications

Our findings support the need for nationwide occupational heat stress standard in the United States. In particular, many of the most burdened states (e.g., Louisiana, Mississippi, Texas) do not have a state Occupational Safety and Health Administration (OSHA) plan to implement a local emphasis program for occupational heat, let alone enact state-level standards.^33^ In the absence of a national standard, employers in these most burdened states are not legally obligated to take provide basic measures (e.g., access to shade, water, and rest) to protect their workers.

Our finding that most socially vulnerable areas are disproportionately burdened by occupational heat exposure is plausible. For example, within the workplace, many heat-exposed workers are paid by the hour or by the piece. These types of payment structures promote over-exertion and, even if permitted by the employer, disincentive workers from taking breaks. Heat-exposed workers often also fall through the cracks in social and labor protections, such as paid leave or rights to unionize, which are critical for promoting health and well-being. The low wages that many heat-exposed workers receive, also, prevent workers from accessing health-promoting assets, such as safe and cool housing or neighborhoods, or adequate preventative care access, which promote body recovery outside of work, and reduce biologic sensitivity to heat. Our findings reinforce the need to broaden social and labor protections for workers. This might be just as important for protecting workers from occupational heat as reducing their heat exposure within the workplace, itself.^34^

Our findings also suggest that occupational health needs to be integrated into climate adaptation policies. Early heat warning systems should target high-risk workers to allow for proactive responses, such as adjusting outdoor work hours.^35^ Health care providers should be trained to identify high-risk workers and better recognize the impacts of work-related heat exposure. Non-occupational social protections, such as healthcare access, should prioritize socially vulnerable working populations.

## Conclusion

The mortality burden of occupational heat exposure is likely far larger than officially reported deaths. Occupational health should be an explicit focus of both work and non-work heat policies, advocacy and adaptation solutions to truly reduce the population health burden of extreme heat.

## Acknowledgements

This publication was supported by the Grant Number, T42 OH008455, funded by the Centers for Disease Control and Prevention. Its contents are solely the responsibility of the authors and do not necessarily represent the official views of the Centers for Disease Control and Prevention or the Department of Health and Human Services. The authors would also like to acknowledge the work by Spangler et al. (daily wet-bulb globe temperature data); Bureau of Labor Statistics (employment count data); Department of Labor (O*NET data); the US CDC National Center for Health Statistics (mortality data); and Kyle Walker (tidycensus R package) for making data feely available for researchers to use.

## Author Contributions

AS and RLN were responsible for study conception and design. AS did the analysis. All authors interpreted the findings and had access to the data. AS and RLN accessed and verified the underlying data. AS wrote the first draft of the manuscript. All authors reviewed and editing the manuscript, approved the current version of the manuscript, and had final responsibility for the decision to submit for publication.

## Data Availability

All data underlying the analysis are publicly available. Yearly county estimates of occupational heat exposure are available at (https://doi.org/10.5281/zenodo.15199025). Major employment counts and sociodemographic compositions are available from the ‘tidycensus’ R package (https://cran.rproject.org/web/packages/tidycensus/index.html). Daily, ambient wet-bulb globe temperatures were previously published by Spangler et al. (https://doi.org/10.6084/m9.figshare.19419836). Metropolitan/nonmetropolitan-level detailed employment counts are available by the Bureau of Labor Statistics Occupational Employment and Wage Statistics (https://www.bls.gov/oes/tables.htm). Workplace characteristics are made publicly available from version 24.1 of the Occupational Information Network (https://www.onetcenter.org/db_releases.html). Mortality data are made publicly available by the CDC National Center for Health Statistics on the WONDER platform (https://wonder.cdc.gov/deaths-by-underlying-cause.html).

## Code Availability

Replication code required to reproduce the results are maintained in the following GitHub repository: *[link to be made available upon publication]*

