## Supplemental Materials for "Mortality burden of outdoor occupational heat exposure in the United States"

**Affiliations:**

**Table of Contents**

**Section 1. Supplementary Tables and Figures for Main Text**

- **Table S1.** Prevalence of occupational heat exposure by year in the contiguous US, 2010-2019.
- **Table S2.** Effect from regression model (per 10% increase in population exposed to occupational heat) and estimated excess occupational heat deaths across all counties, stratifying by 2016 social vulnerability index (SVI) quartile.
- **Table S3.** Composition of employment vs excess occupational heat deaths by social vulnerability quartiles, US region, and race and ethnicity (from Figure 4).
- **Figure S1.** Within-county variability in occupational heat exposure, 2010-2019. Each line represents the difference in the percentage of workers exposed to occupational heat relative to 2010. The orange line reflects the nationwide average trend.
- **Figure S2.** Estimated, annual proportion of all-cause deaths among the working age population (15-64 years old) due to occupational heat exposure by state, averaged across 2010-2019.

**Section 2. Supplemental Methods**

- **Section 2.1. Occupational heat exposure**
  - **Table S4.** Screening criteria using WBGT (in Celsius) for unacclimatized workers set by ACGIH in 2023. From: 2023 TLV®s and BEI®s, © ACGIH 2023.
  - **Table S5.** Counts of detailed SOC groups (n=760) by metabolic rate and work/rest allocation
  - **Table S6.** ACS codes and respective major SOC codes
  - **Figure S3.** Flowchart of how data were harmonized to estimate prevalence of outdoor occupational heat exposure for each year (2010-2019) and geography (county).
  - **Figure S4.** Histogram by detailed SOC group (n=760) of the proportion of work-year outdoors
  - **Figure S5.** Bar chart by detailed SOC group (n=760) of weekend work
  - **Figure S6.** Scatterplot of the previously published rate of occupational heat exposure (per 100 workers, on x-axis) compared to the prevalence of occupational heat exposure estimated in this study (on y-axis).
- **Section 2.2. Sensitivity analyses and robustness checks**
  - **Table S7.** Summary of assumptions and associated checks to assess robustness to assumptions
  - **Table S8.** Sensitivity analysis of main effect from regression model (per 10% increase in population exposed to occupational heat) and estimated excess occupational heat deaths by different approaches to handling suppressed county-year mortality counts
  - **Table S9.** Sensitivity analysis of main effect from regression model (per 10% increase in population exposed to occupational heat) and estimated excess occupational heat deaths by model choice
  - **Table S10.** Sensitivity analysis of occupational heat-mortality effect from regression model (per 10% increase in population exposed to occupational heat) and estimated excess occupational heat deaths after including additional, time-varying covariates
  - **Table S11.** Sensitivity analysis of main effect from regression model (per 10% increase in population exposed to occupational heat) and estimated excess occupational heat deaths by exposure definition
  - **Figure S7.** Full randomization-based placebo test of the estimated occupational heat-mortality relationship. Shaded areas indicate the 0.5th, 1st, 5th, 10th, 25th, 75th, 90th, 95th, 99th, and 99.5th percentiles of the distribution across 500 randomized estimates using completely shuffled exposure prevalences, while the solid black line denotes the median of the randomized estimates. The orange line reproduces our central estimate (shaded orange area is 95% confidence interval) using the true, observed exposure prevalence data, as shown in Fig. 3a.
  - **Figure S8.** Observed vs. fitted deaths from the empirical quasi-Poisson fixed effects model reported in the main text. Each point represents a county x week (n = 30,909). Diagonal orange line denotes 45°.
  - **Figure S9.** Within-county, across year randomization-based placebo test of the estimated occupational heat-mortality relationship. Shaded areas indicate the 0.5th, 1st, 5th, 10th, 25th, 75th, 90th, 95th, 99th, and 99.5th percentiles of the distribution across 500 randomized estimates using completely shuffled exposure prevalences, while the solid black line denotes the median of the randomized estimates. The orange line reproduces our central estimate (shaded orange area is 95% confidence interval) using the true, observed exposure prevalence data, as shown in Fig. 3a.
  - **Figure S10.** Scatterplot of occupational heat exposure prevalence when defined as among all workers (x-axis, as is reported in the main text) instead of the working age population (y-axis).
  - **Figure S11.** Within-year, across county randomization-based placebo test of the estimated occupational heat-mortality relationship. Left panel displays shuffling across all counties in the US, while right panel displays shuffling restricted to a county’s metropolitan-nonmetropolitan area. Shaded areas indicate the 0.5th, 1st, 5th, 10th, 25th, 75th, 90th, 95th, 99th, and 99.5th percentiles of the distribution across 500 randomized estimates using completely shuffled exposure prevalences, while the solid black line denotes the median of the randomized estimates. The orange line reproduces our central estimate (shaded orange area is 95% confidence interval) using the true, observed exposure prevalence data, as shown in Fig. 3a.

**Section 1. Supplemental Tables and Figures for Main Text**

| **Table S1.** Prevalence of occupational heat exposure by year in the contiguous US, 2010-2019 | | | |
| --- | --- | --- | --- |
| **Year(s)** | **Number of workers exposed (millions)** | **Annual average worker count** | **Exposure Prevalence (%)** |
| 2010-2019 | 5.67 (3.76, 8.52) | 148,251,290 | 3.8 (2.5, 5.8) |
| 2010 | 6.87 (4.57, 10.30) | 141,013,149 | 4.9 (3.2, 7.3) |
| 2011 | 5.74 (3.81, 8.64) | 140,881,259 | 4.1 (2.7, 6.1) |
| 2012 | 5.02 (3.33, 7.57) | 142,441,679 | 3.5 (2.3, 5.3) |
| 2013 | 4.59 (3.05, 6.88) | 144,743,387 | 3.2 (2.1, 4.8) |
| 2014 | 4.14 (2.77, 6.19) | 146,982,756 | 2.8 (1.9, 4.2) |
| 2015 | 5.02 (3.34, 7.50) | 149,573,362 | 3.4 (2.2, 5.0) |
| 2016 | 6.48 (4.28, 9.76) | 151,698,102 | 4.3 (2.8, 6.4) |
| 2017 | 5.30 (3.51, 7.98) | 153,814,153 | 3.5 (2.3, 5.2) |
| 2018 | 7.09 (4.68, 10.69) | 154,873,035 | 4.6 (3.0, 6.9) |
| 2019 | 6.64 (4.39, 9.98) | 156,492,017 | 4.3 (2.8, 6.4) |

| **Table S2.** Percent change in working-age all-cause mortality rates (per 10% increase in population exposed to occupational heat) and estimated excess occupational heat deaths across all counties, across years 2010-2019, stratified by quartile based categories of 2016 social vulnerability index (SVI) | | | |
| --- | --- | --- | --- |
| **Model** | **Percent change** | **Relative percent change**** | **Excess deaths***** |
| Overall | 3.3% (1.0, 5.6) | -- | 9,800 (3,100, 17,000) |
| By SVI quartile* | -- | -- | 8,500 (400, 17,400) |
| 1^st^ | 1.4% (-3.1, 5.9) | 0% (reference) | -- |
| 2^nd^ | 2.0% (-1.2, 5.5) | 0.5% (-4.9, 6.2) | -- |
| 3^rd^ | 1.7% (-0.6, 4.5) | 0.3% (-4.7, 5.5) | -- |
| 4^th^ | 4.7% (2.0, 7.5) | 3.3% (-1.9, 8.7) | -- |
| * Higher SVI quartile indicates a higher degree of social vulnerability  ** Relative to 1^st^ SVI quartile  *** Empirically estimated from regression model | | | |

| **Table S3.** Composition of employment vs excess occupational heat deaths by social vulnerability quartiles, US region, and race and ethnicity (from Figure 4). | | | |
| --- | --- | --- | --- |
| **Characteristic** |  | **% of**  **employment** | **% of excess deaths** |
| SVI Quartile | 1^st^ (least vulnerable) | 17.4 | 3.0 |
|  | 2^nd^ | 25.7 | 11.1 |
|  | 3^rd^ | 32.5 | 23.9 |
|  | 4^th^ (most vulnerable) | 24.5 | 62.0 |
| US Region | South | 37.7 | 86.0 |
|  | Midwest | 21.2 | 6.7 |
|  | Northeast | 17.8 | 3.3 |
|  | West | 23.3 | 4.0 |

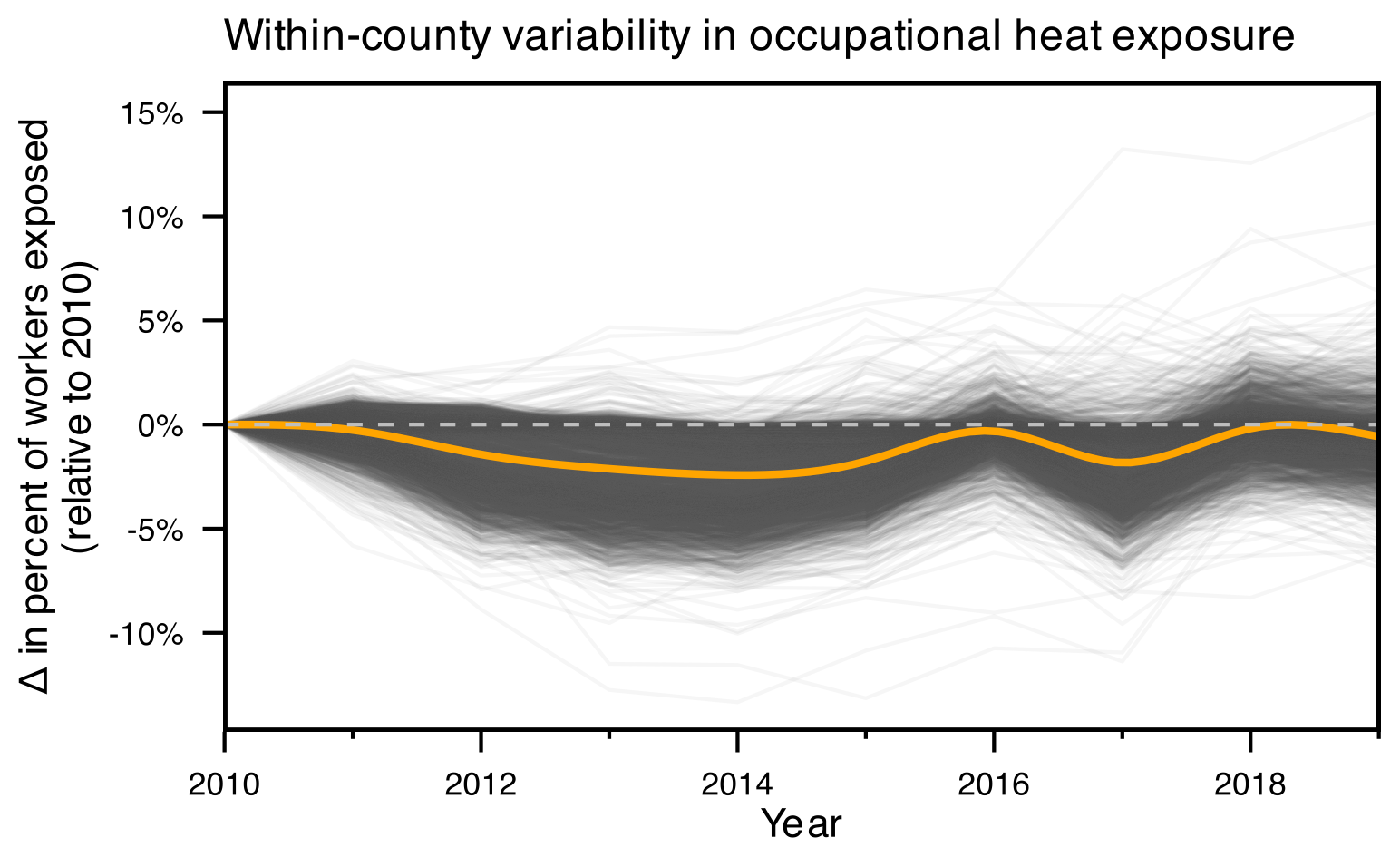

**Figure S1.** Within-county changes in occupational heat exposure, 2010-2019. Each line represents the difference in the percentage of workers exposed to occupational heat relative to 2010. The orange line reflects the nationwide average trend.

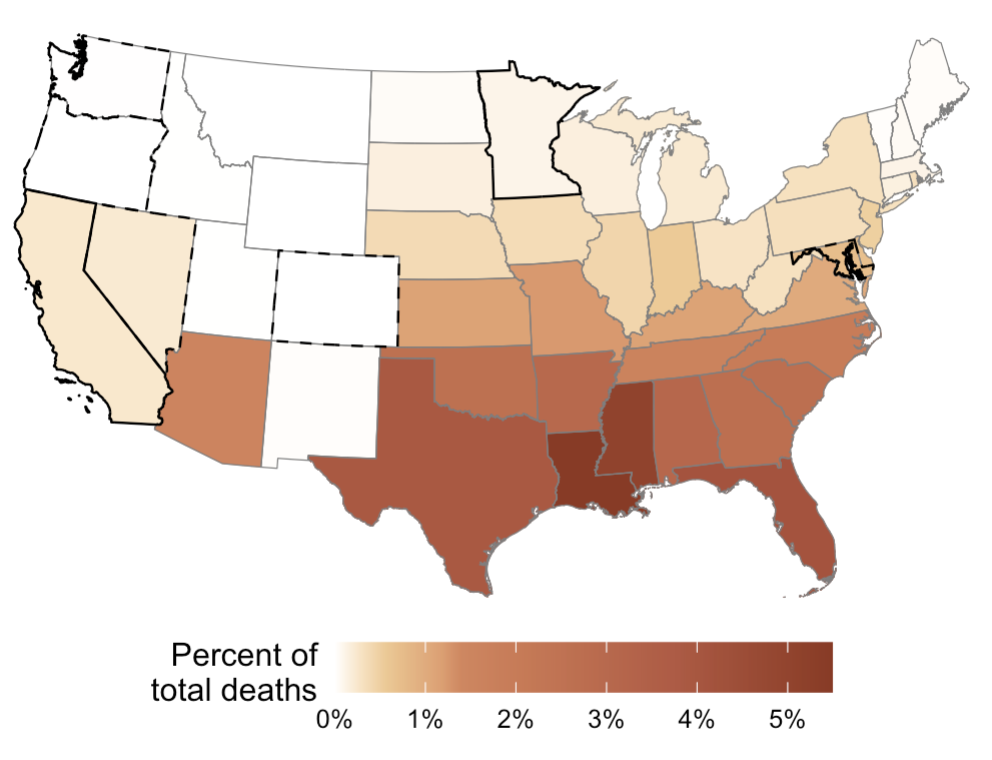

**Figure S2.** Estimated, annual percentage of all-cause deaths among the working age population (15-64 years old) due to occupational heat exposure by state, averaged across 2010-2019.

**Section 2. Supplemental Methods**

**2.1 Occupational heat exposure.** We previously published our methodology and accompanying estimates of occupational heat exposure incidence across the contiguous US during 2010-2019.^1^ Guided by this approach, we re-estimated daily exposure incidence for each county using a more efficient and simplified approach. Below, we provide a detailed methodology to this previously developed approach and highlight how our estimates in this study differ from our previously published estimates.

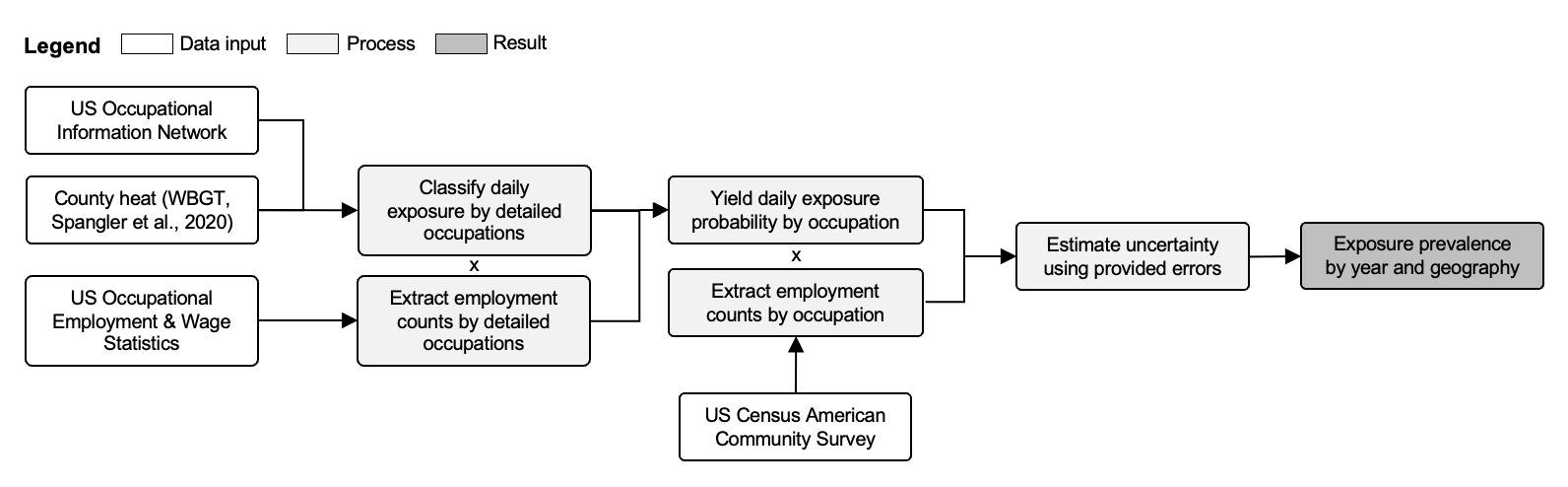

**Figure S3.** Flowchart of how data were harmonized to estimate prevalence of outdoor occupational heat exposure for each year (2010-2019) and geography (county).

**Exposure definition.** We assessed the percentage of all workers likely exposed to ambient heat due to working outdoors. Heat exposure was measured as 24-hr modeled averages of ambient wet-bulb globe temperatures (WBGT; in °C).

**Input data.** Exposure intensity data was extracted from previously modeled, publicly available, county-level, daily wet-bulb globe temperature data during 2010-2019 among the contiguous US (CONUS) only from Spangler et al. (2022).^2^ Exposure frequency, represented as working outdoors, was derived from the US Department of Labor’s Occupational Information Network (O*NET). Typical work characteristics that influence susceptibility to heat exposure were also derived from O*NET. Occupation employment counts by detailed SOC codes were extracted from the US Bureau of Labor Statistics (BLS) Occupational Employment and Wage Statistics (OEWS),^3^ while counts by major SOC codes were extracted from repeated, 5-yr US American Community Survey data.

*Exposure intensity data.* Heat data were estimated by Spangler et al. using raw European Centre for Medium-Range Weather Forecasts (ECMWF) Reanalysis v5 Land product (ERA5-Land) data related to ambient temperature, relative humidity, surface solar radiation downward, wind speed, and surface pressure to calculate WBGT levels using the algorithm developed by Liljegren et al. at a 9 km grid resolution.^4^ This was aggregated to population-weighted, daily, county-level averages. We removed daily data before January 1, 2010 and retained data until December 31, 2019.

We classified each day as “potential exposure” according to 2023 screening criteria thresholds developed by ACGIH,^5^ which are a function of the WBGT levels, the metabolic rate of the worker, and the percentage of time spent working instead of resting. Exposure above these thresholds reflect levels at which an unacclimatized worker may be at risk for heat stress. Thresholds range from 24°C for the workers with least amount of rest and very heavy workload to 30°C for the workers with the most rest and lightest workload.

| **Table S4.** Screening criteria using WBGT (in Celsius) for unacclimatized workers set by ACGIH in 2023. From: *2023 TLV®s and BEI®s, © ACGIH 2023.^5^* | | | | |
| --- | --- | --- | --- | --- |
|  | **Metabolic rate** | | | |
| **Work/rest allocation** | **Light** | **Moderate** | **Heavy** | **Very Heavy** |
| **75 to 100%** | 28 | 25 | *24^a^* | *24^a^* |
| **50 to 75%** | 28.5 | 26 | 24 | *24^a^* |
| **25 to 50%** | 29.5 | 27 | 25.5 | 24.5 |
| **0 to 25%** | 30 | 29 | 28 | 27 |
| ^a^ACGIH does not originally provide screening WBGT values for heavy and very heavy work among those with a high work allocation because of the unsustainability of the work. We set these values to the lowest WBGT value in the table (24°C) | | | | |

*Exposure duration and frequency data.* We leveraged duration and frequency data by detailed, 2010 Standard Occupational Classification (SOC) codes in the 2019 Occupational Information Network (O*NET, version 24.1).^6^ The O*NET database is made publicly available and developed by the US Department of Labor. The O*NET database provides estimates of various work characteristics using self-reported data collected by the Department of Labor. The data are provided by detailed, 2010, O*NET-SOC codes. The 9-digit, detailed O*NET-SOC codes were converted to 7-digit, detailed BLS SOC codes by aggregating the first 7 digits of the O*NET-SOC codes and taking the average value by detailed SOC code.

We determined (i) the probability that a detailed SOC group may be working outdoors, (ii) whether a detailed SOC group typically works long or irregular work schedules, and thus, might work on the weekend, (iii) the metabolic rate of the detailed SOC group, and (iv) allocation of work in a cycle of work and recovery by detailed SOC group.

*Probability of outdoor work.* We used the continuous “Frequency Required to Work Outdoors, Exposed to Weather” work context variable (element ID #4.C.2.a.1.c). Specifically, workers responded to the following question: “How often does this job require working outdoors, exposed to all weather conditions?”. Possible responses ranged from “Never” to “Everyday”. This estimate is provided on a scale of 0-100, which was rescaled to 0-250 to reflect the standard, 250-day work-year, as previously done by our study team. We then converted the estimate back onto a scale of 0-1 to reflect the proportion of the standard work-year that a worker is working outdoors. We replicated this approach for the lower and upper bounds of the provided 95% confidence interval of this variable.

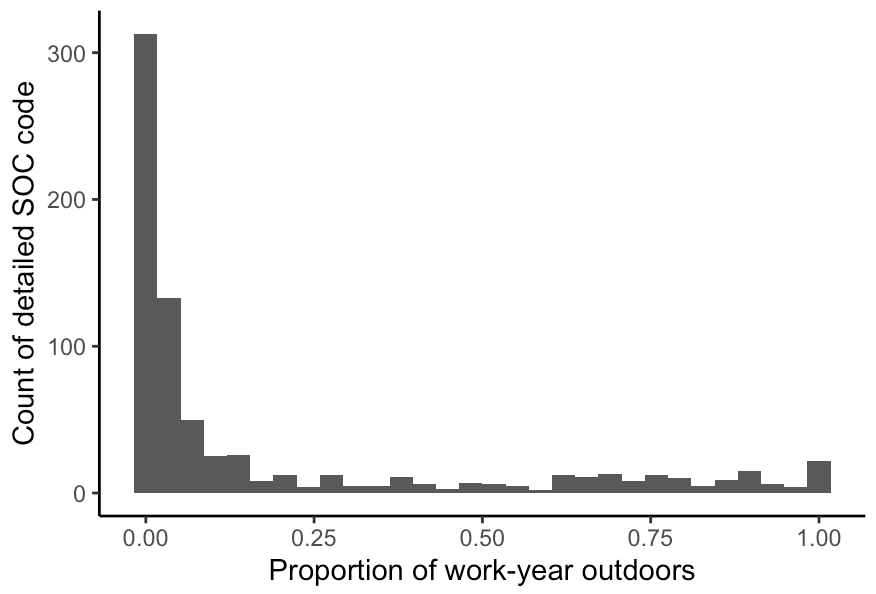

**Figure S4.** Histogram by detailed SOC group (n=760) of the proportion of work-year outdoors

*Long/irregular work schedules.* We used two variables to determine whether a detailed SOC group works long or irregular schedules, and thus, could reasonably work on the weekend. First, we used the continuous “Duration of Typical Work Week” work context variable (element ID #4.C.3.d.8). Specifically, workers responded to the following question: “Number of hours typically worked in one week”. Values ranged from 0-100. Possible responses were from “Less than 40 hours” (value of 0), “40 hours” (value of 50), and “More than 40 hours” (value of 100). We assigned a binary value of whether a detailed SOC group works more than 40 hours if the value was >50. Next, we used the continuous “Work Schedules” work context variable (element ID #4.C.3.d.4). Specifically, workers responded to the following question: “How regular are the work schedules for this job?”. Values ranged from 0-100. Possible responses were from “Regular (established routine, set schedule)” (value of 0), “Irregular (changes with weather conditions, production demands, or contract duration)” (value of 50), and “Seasonal (only during certain times of the year)” (value of 100). We assigned a binary value of whether a detailed SOC group works irregularly if the value was >25. If either the first value was >50 or the second value was >25, we considered the detailed SOC group as likely “weekend workers”.

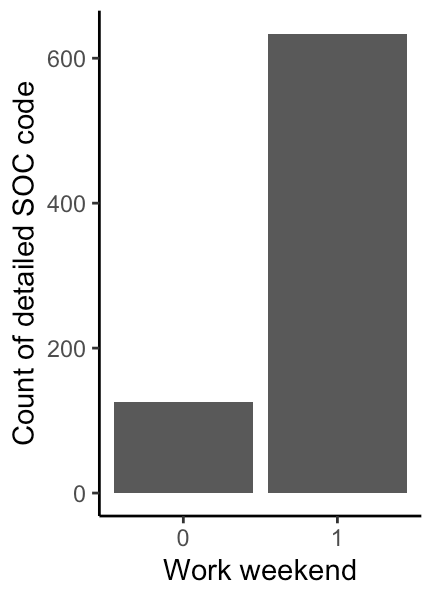

**Figure S5.** Bar chart by detailed SOC group (n=760) of weekend work

*Metabolic rate.* We used four variables to determine metabolic rate. We assigned each detailed SOC group as “light”, “moderate”, “heavy”, or “very heavy” metabolic rate if one of the (i) time spent sitting, (ii) time spent standing, (iii) average time spent standing and using hands, and (iv) time spent walking/running has the highest possible value, respectively.

First, we used the continuous “Time Spent in Body Positions, Spend Time Sitting” work context variable (#4.C.2.d.1.a) for (i) light metabolic rate. Specifically, workers answered “How much does this job require sitting?”. Second, we used the continuous “Time Spent in Body Positions, Spend Time Standing” work context variable (#4.C.2.d.1.b) for (ii) moderate metabolic rate. Specifically, workers answered “How much does this job require standing?”. Third, we took the average of this previous variable and the continuous “Time Spent in Body Positions, Spend Time Using Your Hands to Handle, Control, or Feel Objects, Tools, or Controls” work context variable (#4.C.2.d.1.g) for (iii) heavy metabolic rate. Specifically, workers answered “How much does this job require using your hands to handle, control, or feel objects, tools or controls?”. Fourth, we used the continuous “Time Spent in Body Positions, Spend Time Walking and Running” work context variable (#4.C.2.d.1.d) for (ii) very heavy metabolic rate. Specifically, workers answered “How much does this job require walking and running?”. Each of the four factors had values ranging from 0-100 (from “Never” to “Continually or almost continually”). The factor that had the highest value among a detailed SOC group was assigned as the metabolic rate for that group.

*Work/rest allocation.* To determine the work vs recovery time, we again used the continuous “Time Spent in Body Positions, Spend Time Sitting” work context variable (#4.C.2.d.1.a). We considered time spent sitting as “recovery time”, and time spent not sitting as “work time”. Below is a table that breaks down the spread of 760 detailed SOC groups by metabolic rate and work/rest allocation.

| **Table S5.** Counts of detailed SOC groups (n=760) by metabolic rate and work/rest allocation | | | | |
| --- | --- | --- | --- | --- |
|  | **Metabolic rate** | | | |
| **Work/rest allocation** | **Light** | **Moderate** | **Heavy** | **Very Heavy** |
| **75 to 100%** | 0 | 101 | 76 | 3 |
| **50 to 75%** | 2 | 79 | 100 | 6 |
| **25 to 50%** | 160 | 8 | 45 | 1 |
| **0 to 25%** | 179 | 0 | 0 | 0 |

*Employment counts.* For county-level, detailed SOC codes, we extracted 2019 metropolitan/nonmetropolitan area employment counts from the US Bureau of Labor Statistics (BLS) Occupational Employment and Wage Statistics (OEWS).^3^ Metropolitan areas are counties grouped together based on common economic activities and commuting patterns around a city. Thus, some metropolitan areas are a single county, while others are a combination of a few counties. Nonmetropolitan areas are larger groups of many counties that do not have economic activity around a major city. We assumed that the spread of detailed SOC codes was the same for the counties within a metro/nonmetro area. These detailed SOC employment counts are not developed to be assessed in a time-varying manner, as the OEWS aggregates data across many years to develop their estimates. Thus, we used 2019 data to span across the years of analysis in this study (2010-2019). In 2018, SOC codes were updated from the 2010 SOC code version. We integrated all data to 2010 SOC codes since the majority of data used in this study used the 2010 SOC structure. Thus, we crosswalked 2018 detailed SOC codes from the OEWS to 2010 detailed SOC codes.

For county-level major SOC codes, the 5-yr US American Community Survey provides employment counts by 2-digit, 2012 NAICS codes for every county. These employment counts by sex were extracted using the `tidycensus` R package and the `get_acs()` function from Table C24030 (“Sex by Industry for the Civilian Population”). The specific ACS variable codes are displayed in **Table S6**. The sex-specific estimates were aggregated by county.

| **Table S6.** ACS codes and respective major SOC codes | | | |
| --- | --- | --- | --- |
| ACS variable | |  |  |
| Male | Female | SOC | Occupation |
| C24010_005 | C24010_041 | 11-0000 | Management |
| C24010_006 | C24010_042 | 13-0000 | Business and Financial Operations |
| C24010_008 | C24010_044 | 15-0000 | Computer and Mathematical |
| C24010_009 | C24010_045 | 17-0000 | Architecture and Engineering |
| C24010_010 | C24010_046 | 19-0000 | Life, Physical, and Social Science |
| C24010_012 | C24010_048 | 21-0000 | Community and Social Service |
| C24010_013 | C24010_049 | 23-0000 | Legal |
| C24010_014 | C24010_050 | 25-0000 | Education, Training, and Library |
| C24010_015 | C24010_051 | 27-0000 | Arts, Design, Entertainment, Sports, and Media |
| C24010_016 | C24010_052 | 29-0000 | Healthcare Practitioners and Technical |
| C24010_020 | C24010_056 | 31-0000 | Healthcare Support |
| C24010_021 | C24010_057 | 33-0000 | Protective Service |
| C24010_024 | C24010_060 | 35-0000 | Food Preparation and Serving Related |
| C24010_025 | C24010_061 | 37-0000 | Building and Grounds Cleaning and Maintenance |
| C24010_026 | C24010_062 | 39-0000 | Personal Care and Service |
| C24010_028 | C24010_064 | 41-0000 | Sales and Related |
| C24010_029 | C24010_065 | 43-0000 | Office and Administrative Support |
| C24010_031 | C24010_067 | 45-0000 | Farming, Fishing, and Forestry |
| C24010_032 | C24010_068 | 47-0000 | Construction and Extraction |
| C24010_033 | C24010_069 | 49-0000 | Installation, Maintenance, and Repair |
| C24010_035 | C24010_071 | 51-0000 | Production |
| C24010_036 | C24010_072 | 53-0000 | Transportation and Material Moving |
| C24010_037 | C24010_073 | 53-0000 | Transportation and Material Moving |

**Estimation approach.** We directly estimated the proportion of workers ($p_{dco_{1}}$) exposed to outdoor heat exposure for every $d$ day from 2010-2019, broken down by $c$ county and $o_{1}$ major SOC group:

$$p_{dco_{1}}=\frac{\sum_{o_{2}=1}^{n} ({Outdoor}_{o_{2}}\times1({{WBGT}_{dc}>T}_{o_{2}})\times N_{co_{2}})}{\sum_{o_{2}=1}^{n} N_{co_{2}}}$$

where $o_{2}$ reflects the $n$ detailed SOC groups within a given major SOC group; ${Outdoor}_{o_{2}}$ is the estimated probability of working outdoors for a given detailed SOC group; $1({{WBGT}_{dc}>T}_{o_{2}})$ is an indicator variable for whether the ambient wet-bulb globe temperature levels on a given county-day exceed the threshold of exposure for a given detailed SOC group (**Table S4**); and $N_{co_{2}}$ reflects the total number of workers in a particular detailed SOC group within a particular county.

**Post-stratification.** The estimated proportion of workers exposed ($p_{dco_{1}}$) to outdoor heat for each of the $o_{1}$ major SOC groups for each $d$ day within $c$ county during 2010-2019 can be used to estimate the daily percentage of all workers exposed for $c$ county ($\hat{P}_{dc}$):

$$\hat{P}_{dc}={\sum_{o_{1}=1}^{22} (p_{dco_{1}}*{Pop}_{cyo_{1}})}/{{Pop}_{cy}}$$

where ${Pop}_{cyo_{1}}$ and ${Pop}_{cy}$ represent the county-level working population counts for each year in a specific major SOC group or all workers, respectively. To estimate uncertainty, we used the lower and upper limits of the probability of working outdoors for a given detailed SOC (${Outdoor}_{o_{2}}$) to estimate an empirical 95% confidence interval for each $\hat{P}_{dc}$.

To estimate yearly prevalence while considering that the same workers could be exposed to heat on different days, we adapt and applied a previously developed method^7^ to estimate the overall exposure prevalence ($P_{cy}$) for county $c$ in year $y$ when the variation in between-day exposure incidence is dependent:

$$\hat{P}_{cy}=1-\prod_{d=1}^{365} [1-(1-\rho)\times\hat{P}_{cd}]$$

where $d$ is the number of days in a year (366 for leap-years); $\hat{P}_{cd}$ is the proportion of workers exposed on day $d$ within county $c$; and $\rho$ reflects the correlation in the number of workers exposed between each day. The true between-day exposure correlation is unknown, so we conservatively specify a time-invariant $\rho=\text{0.99}$ to reflect that the workers exposed to any outdoor hazard each day are highly similar, but not always the same (i.e, $\rho=\text{1}$).

**Output.** For every county in the US during 2010-2019, we estimated the annual proportion of workers exposed to outdoor occupational heat. Our approach allows for convenient aggregation of state-, region-, and nationwide-specific prevalence estimates.

**Difference from previous estimation methodology.** A key component of our previously published methodology that made our estimation less computationally efficient than the estimates presented in this study was the estimation of the proportion of workers who were likely to be working outdoors on any given day as derived from the O*NET.

Using this proportion as a probability, we previously published our estimates using Monte Carlo simulation with 200 iterations to estimate how many workers in a particular detailed SOC group were likely to be working outdoors on a particular day. This meant that we ran 200 iterations x 365 days x 10 years = 730,000 simulations. However, with enough Monte Carlo simulations, the central estimate from this approach would be equivalent to simply multiplying the outdoor work probability with the number of workers in a particular detailed SOC code. This modified approach is substantially more computationally efficient for a similar result. To estimate uncertainty in the outdoor work probability, we instead used standard errors provided by O*NET to generate 95% confidence intervals to our estimate.

**Figure S6** shows the relationship between the exposure rates per 100 workers as previously published in Shkembi et al. (2025) using the Monte Carlo simulation approach compared to the constructed prevalence estimates in this study. The two indicators were highly correlated (Pearson’s ρ = 0.994), indicating that our updated approach to estimate prevalence did not artificially introduce substantial bias to our occupational heat exposure estimates.

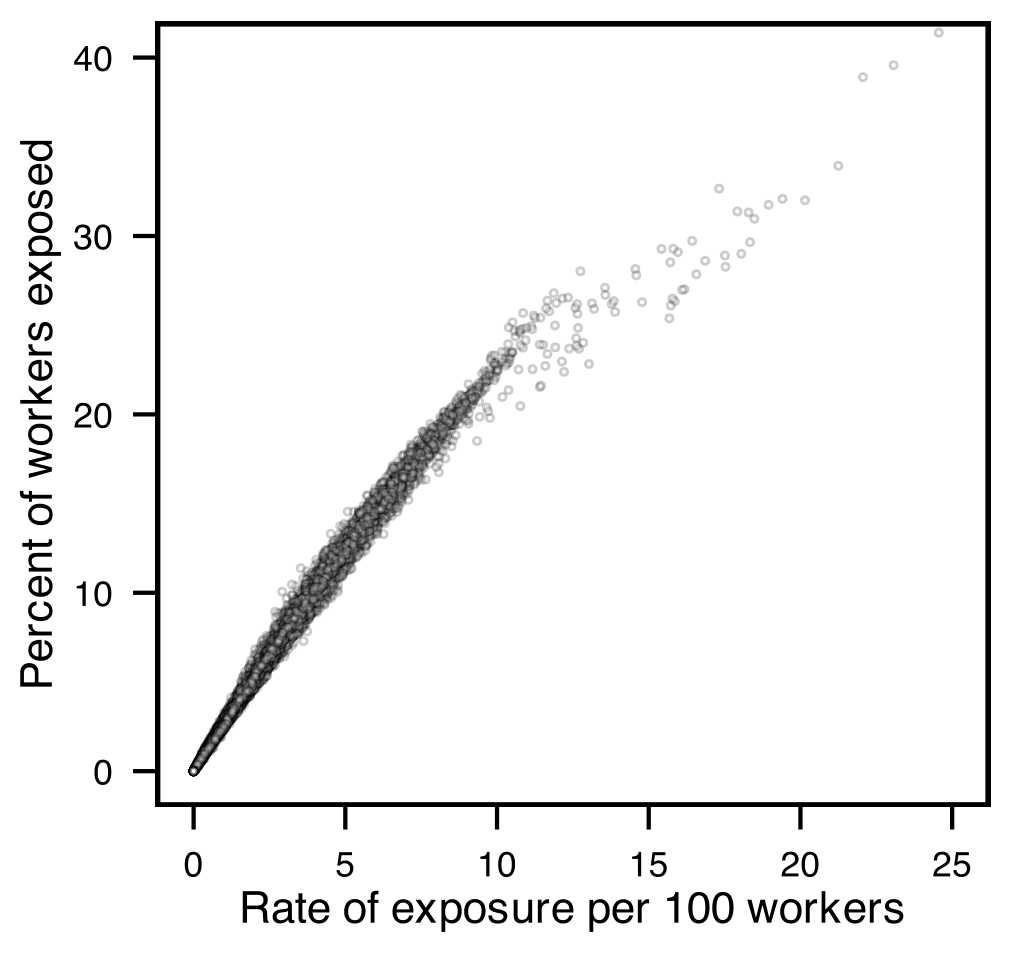

**Figure S6.** Scatterplot of the previously published rate of occupational heat exposure (per 100 workers, on x-axis) compared to the prevalence of occupational heat exposure estimated in this study (on y-axis).

**2.2 Sensitivity analyses and robustness checks.** Given the absence of any prior analyses that estimate the full mortality burden of occupational heat, it is not possible to subjectively compare our estimates to previous analyses. Thus, there is a risk of observing spurious effects as a result of violating various assumptions related to causal inference.^8^ Below, we describe a series of sensitivity analyses and robustness checks that test our ability to observe a causal effect of occupational heat on mortality. These checks are summarized in **Table S7**. Overall, we do not find significant evidence that indicates we have violated these assumptions, suggesting that we have identified causal effects between the occupational heat and mortality.

| **Table S7.** Summary of assumptions and associated checks to assess robustness to assumptions | |
| --- | --- |
| **Assumption** | **Test, check, or analysis** |
| Valid data quality | *Suppressed mortality data:* Examine model sensitivity to different approaches to handling suppressed mortality counts for county-years with <10 deaths |
|  | *Placebo test for occupational heat exposure:* Scramble exposure prevalence across both years and counties |
| Regression is correctly specified | *Regression model choice:* Use negative binomial regression model instead of quasi-Poisson |
|  | *Model fit:* Assess observed vs. fitted values |
| Exchangeability | *Placebo test for unmeasured time-invariant confounding:* Scramble exposure prevalence across years within each county |
|  | *Additionally measured time-varying confounding:* Control for educational attainment, income, employment status, healthcare access, air quality, and extreme cold |
|  | *Potential effect of unmeasured time-varying confounding:* Estimate e-value |
| Consistency | *Exposure definition:* Define exposure prevalence among the entire working age population rather than among workers only |
|  | *No cross-county spillover effects:* Scramble exposure prevalence across counties within each year |

**Valid data quality.** Our ability to observe unbiased occupational heat-mortality effects relies on the assumption that our exposure and outcome adequately reflect what they were intended to measure.

*Suppressed mortality data.* We extracted publicly available mortality counts for 3,108 contiguous US counties between 2010-2019 from the National Center for Health Statistics.^9^ The mortality counts were broken down by county and year, and counts included deaths from all-causes for all ages. For simplicity, we chose to use publicly available mortality data rather than the restricted, full data since it has been shown that using either of the two data sources when using a similar statistical methodology (i.e., quasi-Poisson panel fixed effects modeling) in the context of environmental exposures will yield similar model coefficients.^10^ However, for some county-years (8% of all county-years), mortality counts were suppressed due to a low number of deaths (<10 deaths). In the model results reported in the main text, we imputed the midpoint of 0-9 deaths (i.e., 4.5 deaths) for each suppressed county-year value to obtain the largest sample size during modeling. We conducted additional sensitivity analyses to explore whether the model results are altered by different choices of handling the suppressed values: (1) impute 0, (2) impute 9, or (3) drop suppressed values. **Supplemental Table S8** displays that the main effect of occupational heat exposure on all-cause mortality is not qualitatively altered by any of these choices.

| **Table S8.** Sensitivity analysis of main effect from regression model (per 10% increase in population exposed to occupational heat) and estimated excess occupational heat deaths by different approaches to handling suppressed county-year mortality counts | | |
| --- | --- | --- |
| **Approach** | **Model coefficient** | **Excess deaths** |
| Impute 4.5 | 3.3% (1.0, 5.6) | 9,800 (3,100, 17,000) |
| Impute 0 | 3.2% (0.9, 5.6) | 9,500 (3,100, 16,800) |
| Impute 9 | 3.4% (1.1, 5.7) | 10,100 (3,200, 17,100) |
| Drop | 3.4% (1.2, 5.8) | 10,300 (3,400, 17,500) |

*Placebo test for occupational heat exposure estimation.* Our exposure prevalence estimates were constructed with the intent of accurately measuring potential exposure occupational heat. While our approach is theoretically valid, we previously identified limitations that can bias estimation of exposure in a given county.^1^ Thus, we additionally used a randomization-based placebo test to investigate the quality of our exposure prevalence estimates.^11^ If we shuffle the exposure prevalence estimates across both counties and years, we should break any association between exposure prevalence and mortality. This randomization should yield a null effect estimate, also known as a negative exposure control.^12^ If the estimated effect is not null, the placebo test would suggest that our modeled effects are being driven by potentially spurious correlations due to mismeasured or non-valid exposure prevalence estimates. We scrambled estimated exposure prevalence 500 times and reran the unstratified quasi-Poisson model as specified in the main text (Eq. 2). **Figure S7** shows the estimated effect of occupational heat exposure prevalence on mortality using shuffled exposure prevalence, compared to our observed estimates. The results suggest that the effect of nonvalid exposure prevalence estimates is minimal, as we observe a null effect on average when exposure prevalences are fully shuffled across counties and years.

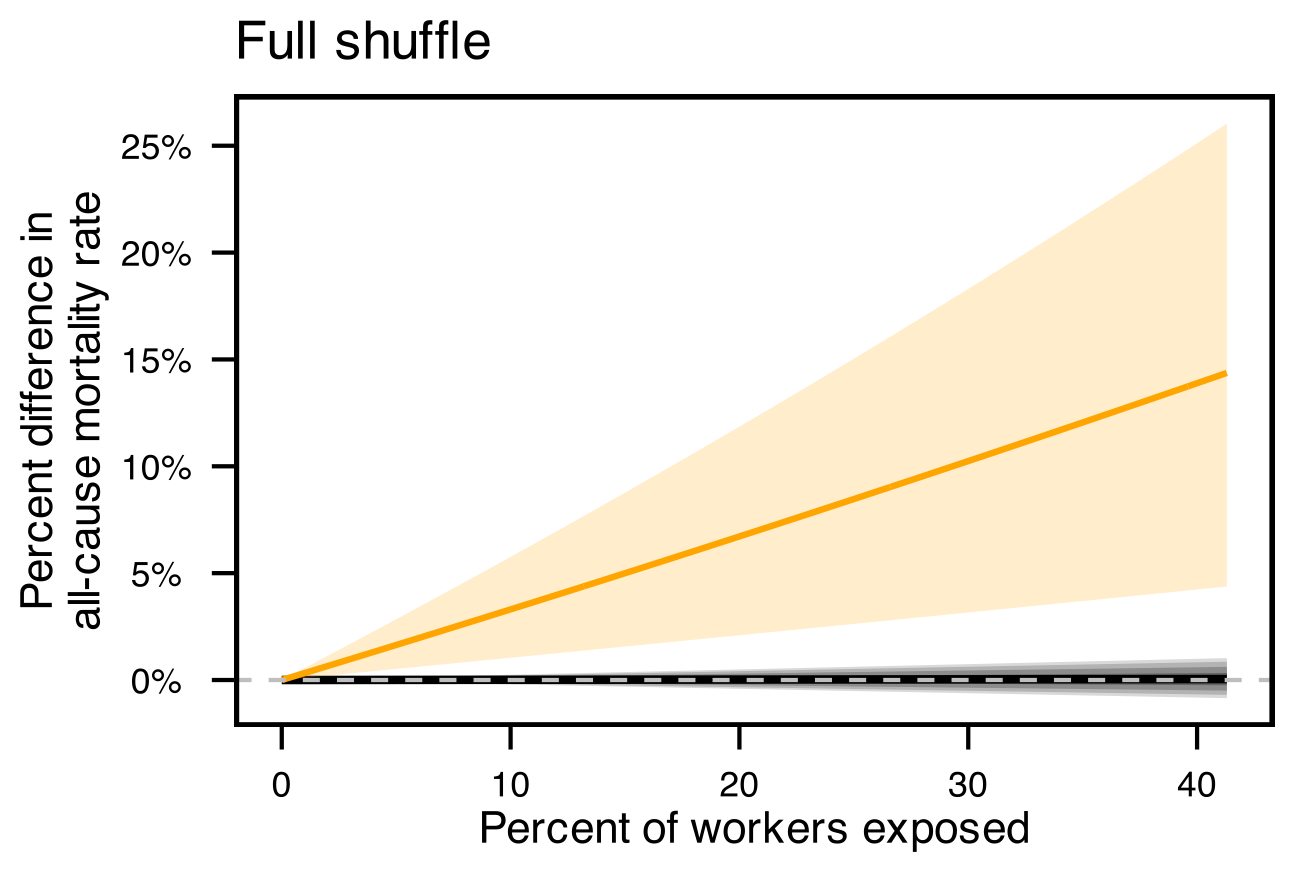

**Figure S7.** Full randomization-based placebo test of the estimated occupational heat-mortality relationship. Shaded areas indicate the 2.5th, 5th, 10th, 25th, 75th, 90th, 95th, and 97.5th percentiles of the distribution across 500 randomized estimates using completely shuffled exposure prevalences, while the solid black line denotes the median of the randomized estimates. The orange line reproduces our central estimate (shaded orange area is 95% confidence interval) using the true, observed exposure prevalence data, as shown in **Fig. 3a**.

**Regression is correctly specified.** A key assumption of observing a causal effect is that the model used to generate the effect is correctly specified. We examine whether our results are robust to choice of regression and examine the fit of our main model to probe at this assumption.

*Regression model choice.* While our choice of quasi-Poisson regression is theoretically valid to account for the mortality count outcome data, overdispersion in mortality counts, and between-county differences in the total population (via offset), negative binomial regression could be theoretically appropriate as well. We re-analyzed the estimated occupational heat-mortality effect using a negative binomial model to ensure that our observed effects are not driven by the choice of a quasi-Poisson model. The negative binomial model included the same outcome, covariates, and offset as the model reported in the main text. **Table S9** indicates that the quasi-Poisson regression model is qualitatively similar to the negative binomial model.

| **Table S9.** Sensitivity analysis of main effect from regression model (per 10% increase in population exposed to occupational heat) and estimated excess occupational heat deaths model choice | | |
| --- | --- | --- |
| **Approach** | **Model coefficient** | **Excess deaths** |
| Quasi-Poisson | 3.3% (1.0, 5.6) | 9,800 (3,100, 17,000) |
| Negative Binomial | 3.1% (1.0, 5.4) | 9,400 (2,900, 16,400) |

*Model fit*. We assessed the goodness of fit of the quasi-Poisson fixed effects model reported in the main text using pseudo-R^2^ values and plotting the observed vs. fitted mortality relationship (**Figure S8**). The model exhibited good fit (pseudo-R^2^ = 0.999) and predicted the observed mortality counts well, indicating that our model adequately captures the variability in mortality.

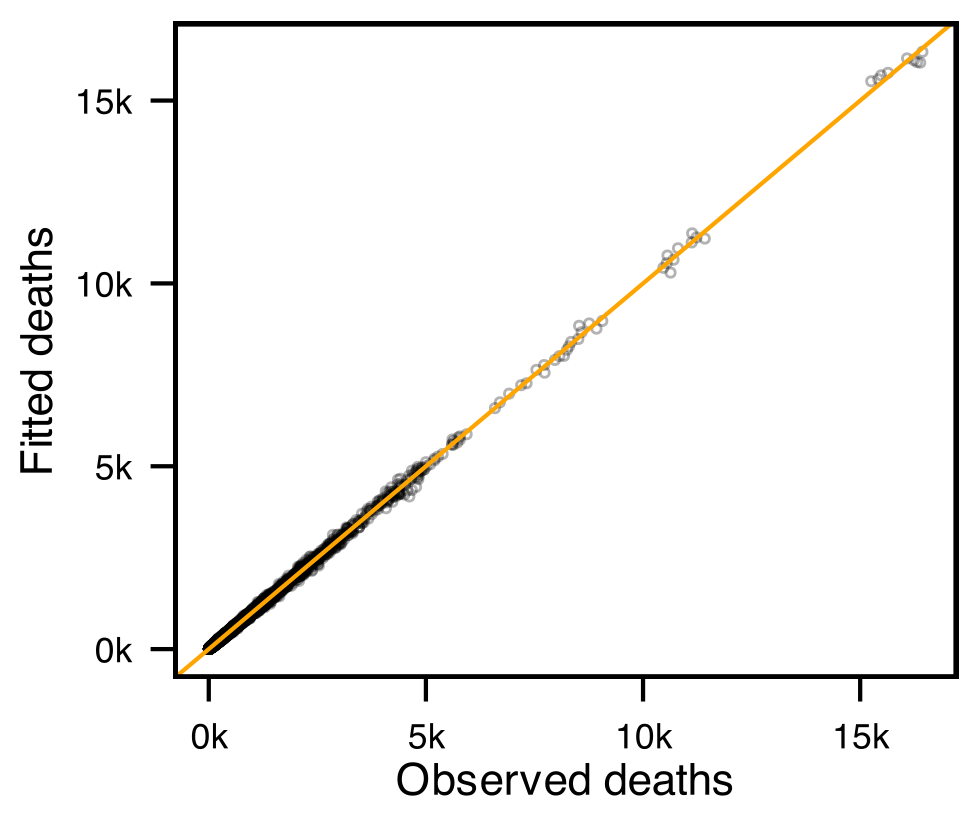

**Figure S8.** Observed vs. fitted deaths from the empirical quasi-Poisson fixed effects model reported in the main text. Each point represents a county-year. Diagonal orange line denotes 45°.

**4.3 Exchangeability.** An important assumption of causal inference is that of exchangeability, which can be achieved by randomly assigning exposures. It is well understood, however, that randomization of occupational risks is neither plausible nor ethical.^13^ Occupational epidemiology instead relies on carefully selecting potential confounders using observational data. Our model approach here accounts for any time-invariant confounders between counties and accounts for a robust set of time-varying confounders but relies on the assumption that there are no additional unmeasured confounders. We conducted three sensitivity analyses and tests to probe at this assumption. Together, these three checks provide further evidence that our findings are unlikely to be driven by unmeasured confounding.

*Placebo test for unmeasured time-invariant confounding.* The county fixed effect covariate theoretically accounts for any county-specific, time-invariant factors that could be associated with both occupational heat exposure and mortality, such as rurality. To test this assumption, we conducted an additional within-county randomization placebo test that shuffles exposure prevalence over time.^11^ The exposure prevalence is always assigned to the correct county, but the year of exposure is random. By breaking the temporal structure of the exposure prevalence, if we still observe an effect between occupational heat and mortality, then the effect could be driven by a time-invariant confounder between counties that might be generating spurious associations. For each occupational risk factor, we scrambled estimated exposure prevalence within each county 500 times and reran the quasi-Poisson model as specified in the main text (Eq. 2). **Figure S9** shows the estimated effect of occupational heat exposure prevalence on mortality using the within-county shuffled exposure prevalence, compared to our observed estimates. The results suggest that confounding bias from an unmeasured time-invariant factor is minimal, as we observe a null effect on average when exposure prevalences are shuffled across years and within counties.

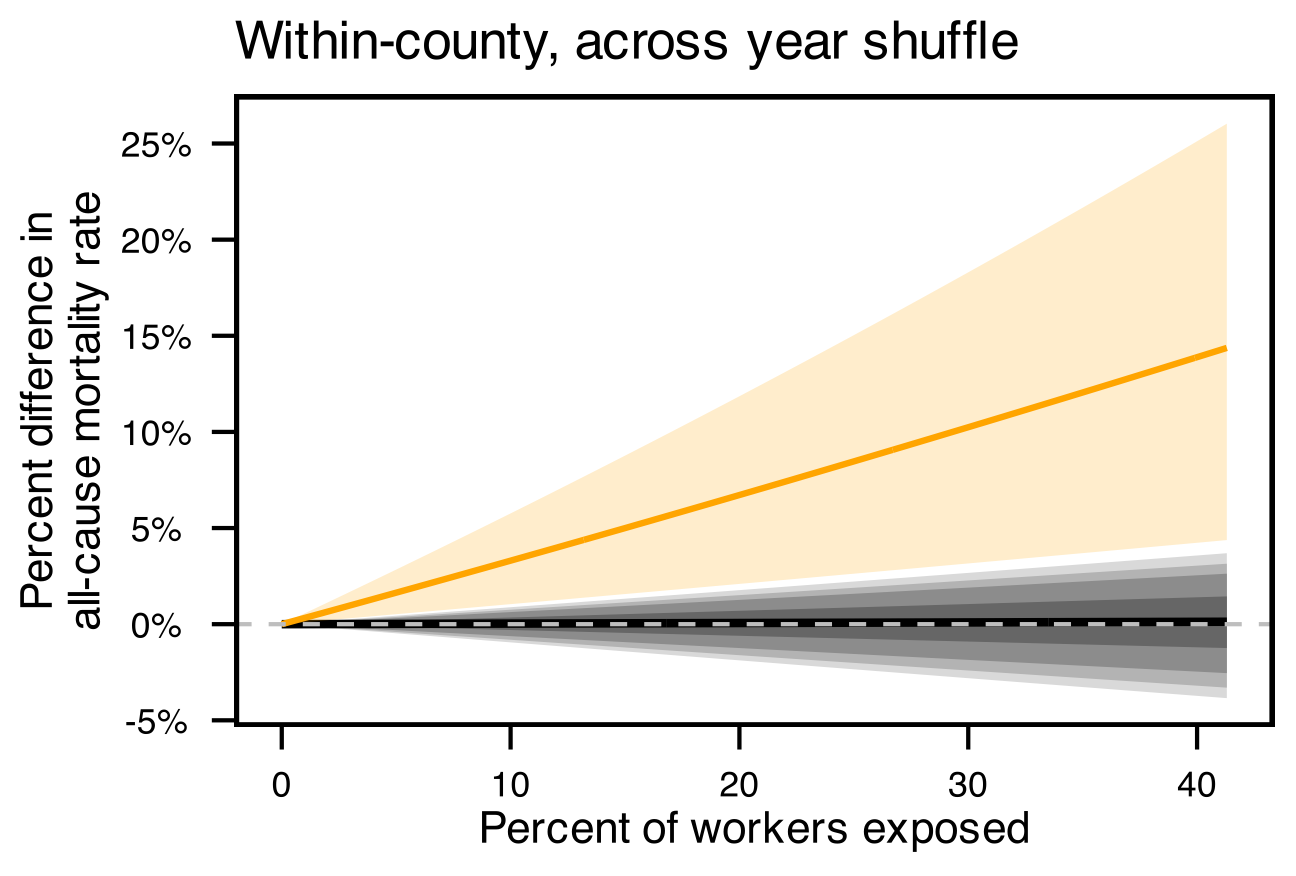

**Figure S9.** Within-county, across year randomization-based placebo test of the estimated occupational heat-mortality relationship. Shaded areas indicate the 2.5th, 5th, 10th, 25th, 75th, 90th, 95th, and 97.5th percentiles of the distribution across 500 randomized estimates using completely shuffled exposure prevalences, while the solid black line denotes the median of the randomized estimates. The orange line reproduces our central estimate (shaded orange area is 95% confidence interval) using the true, observed exposure prevalence data, as shown in **Fig. 3a**.

*Additionally measured time-varying confounding.* While we minimized over-adjustment of potential time-varying factors that are plausibly downstream of occupational heat exposure, some factors may reasonably mediate or confound the relationship. These could include educational attainment, income, employment status, health insurance status, air quality, and extreme cold exposure. Given our conceptual diagram, we determined that many of these factors are likely downstream of occupational heat exposure and did not include such factors as model covariates since our goal was to assess the full mortality burden of occupational heat exposure. Here, we examined the estimated occupational heat-mortality effect after additionally controlling for these time-varying factors.

Educational attainment was measured as the yearly percentage of residents >25 years old in a county who do not have a high school diploma from repeated, 5-yr US American Community Survey (ACS) data during 2010-2019. Specifically, we used variables “002”-“016” from Table B15003 to reflect those without a high school diploma, and variable “001” to reflect the number of residents aged >25 years old. Income was measured as the yearly percentage of residents in a county who are below 200% of the Federal Poverty Limit from repeated, 5-yr US ACS data during 2010-2019. Specifically, subtracted variable “008” (those above 200% of the Federal Poverty Limit) from “001” (the total number of residents) from Table C17002. Employment status was measured as a yearly percentage of residents in a county who are unemployed, do not partake in the labor force, or are in the military from repeated, 5-yr US ACS data during 2010-2019. Specifically, we used variables “005” (unemployed), “006” (in armed forces), and “007” (not in labor force) from Table B23025 in the ACS. Health insurance status was measured as the yearly percentage of the employed residents of a county who do not have health insurance from repeated 5-yr US ACS data during 2010-2019. Specifically, we used variables “008” (18-64 years old) and “013” (65+ years old) between 2010-2014 and variable “007” between 2015-2019 to reflect those without health insurance, and variable “003” to reflect the number of employed residents from Table B27011.

Air quality was measured as the county-level, annual average, ambient concentration of fine particulate matter (PM_2.5_; µg/m³), nitrogen dioxide (NO2; ppb), and sulfur dioxide (SO2; ppb) from 2010 to 2019 as estimated by the Center for Air, Climate, and Energy Solutions (CACES) land use regression model.^14,15^ We additionally estimated the annual number of extreme cold days using a similar approach to estimating the number of extreme heat days in the main text. For each county-year, extreme cold was defined as days with daily wet-bulb globe temperature lower than the county’s 10th percentile cold season temperature (November to March, 2010 to 2019) and the days were summed.^2^ The 10^th^ percentile threshold was arbitrarily selected to reflect exposure to relatively extreme cold days within a county.

We included these county-year variables as covariates of our main quasi-Poisson model, both individually and collectively, to compare the estimated occupational heat-mortality effect with and without the additional, time-varying covariates. **Table S10**). indicates that our estimated occupational heat-mortality effects and their associated excess deaths are qualitatively similar when including any of these potential time-varying confounders as covariates. When all potential confounders are included, the estimated annual excess deaths was even larger (11,700 vs 9,800). This suggests that our findings are likely robust to a large set of measurable, potential time-varying confounders.

| **Table S10.** Sensitivity analysis of occupational heat-mortality effect from regression model (per 10% increase in population exposed to occupational heat) and estimated excess occupational heat deaths after including additional, time-varying covariates | | |
| --- | --- | --- |
| **Approach** | **Model coefficient** | **Excess deaths** |
| Main model | 3.3% (1.0, 5.6) | 9,800 (3,100, 17,000) |
| + no HS diploma | 3.3% (1.1, 5.7) | 9,700 (3,100, 17,000) |
| + low-income | 3.2% (1.0, 5.5) | 9,400 (3,000, 16,600) |
| + not working | 3.5% (1.3, 5.8) | 10,400 (4,000, 17,600) |
| + uninsured | 3.2% (1.0, 5.6) | 9,700 (3,100, 17,000) |
| + PM_2.5_ | 3.5% (1.3, 5.8) | 10,500 (3,700, 17,600) |
| + NO_2_ | 3.4% (1.2, 5.7) | 10,200 (3,600, 17,400) |
| + SO_2_ | 3.0% (0.8, 5.4) | 9,000 (2,400, 16,200) |
| + extreme cold | 3.2% (1.0, 5.7) | 9,700 (3,000, 17,200) |
| + all covariates | 3.9% (1.7, 6.1) | 11,700 (5,100, 18,500) |

*Potential effect of unmeasured time-varying confounding.* While it is impossible to determine whether any potential confounders remain unmeasured, E-values can be useful to examine the minimum strength of association that an unmeasured confounder would need to have with the occupational heat and mortality, conditional on the measured confounders, to fully explain away the observed effect.^16^ We calculated an E-value for ${RR}_{1}=\exp\left( \beta_{1} \right)=1.0034$ using the following equation:^16^

$${E\text{-}value}_{1}={RR}_{1}+ \surd({RR}_{1}\times({RR}_{1}-1))$$

which estimates an ${E\text{-}value}_{1}$ = 1.06. This suggests that the observed percent change in mortality rate from a 1% increase in the percentage of workers exposed to occupational heat could be explained away by an unmeasured confounder that was associated with both occupational heat and mortality by an RR of 1.06, conditional on any time-invariant, between-county confounders and time-varying confounding state-level policies and due to heat and precipitation. Should such unmeasured confounder exist, it would likely be related to lifestyle factors (e.g., poor diet, smoking, alcohol consumption), which have been shown to be associated with more hazardous employment.^17,18^ However, much, if not all, of the confounding effect due to these unmeasured lifestyle factors is likely already accounted by the county fixed effects (which would capture time-invariant differences between counties due to characteristics such as rurality and food access). Additionally adjusted for time-varying SES factors (income, education, labor force participation, healthcare insurance status, ambient air quality, as displayed in the previous subsection) did not qualitatively alter our estimated effect either, and these SES factors are also associated with lifestyle factors.^19^ This suggests it is unlikely that an unmeasured, time-varying confounder could entirely explain our observed effect.

**4.4 Consistency.** Another assumption of causal inference is consistency, which can be achieved through clear and well-defined exposure information. Our approach to characterize occupational heat as the percentage of workers who are likely to be exposed to wet-bulb globe temperatures high enough to cause heat stress among unacclimated workers theoretically satisfies this assumption. We could have defined exposure prevalence among the entire working age population (including individuals who are unemployed or not in the labor force), instead of only among employed workers. We reran our main quasi-Poisson model using this alternative exposure prevalence definition to investigate whether our exposure definition produces a spurious effect. Another manner that the consistency assumption can be violated is if the potential mortality outcome of one county is affected by occupational heat exposure from another county. Such spillover effect could be possible due to shared economic patterns between some counties, wherein a worker resides in one county but works in a neighboring county. We also conducted two across-county randomization placebo tests to investigate whether our estimated effects for each occupational risk factor could be driven by spillover effects across counties.^11^

*Exposure definition.* Defining exposure prevalence among the working age population is highly correlated with defining it among workers only (Pearson’s ρ = 0.982; **Figure S10**). Rerunning our quasi-Poisson regression model defining exposure prevalence among the entire working age population produced a similar effect on all-cause mortality, but was less precise (**Table S11**). The estimated excess deaths were smaller (7,900 vs 9,800 in the main text), but within the range of uncertainty.

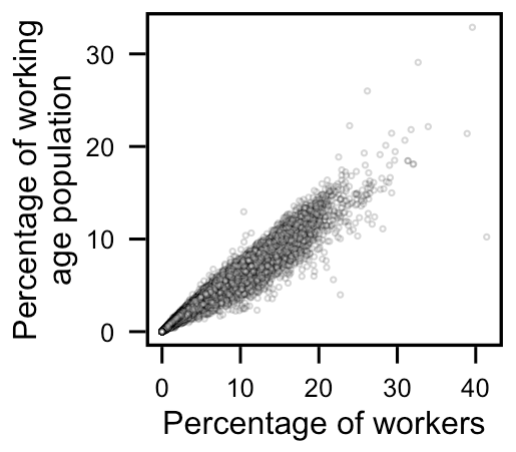

**Figure S10.** Scatterplot of occupational heat exposure prevalence when defined as among all workers (x-axis, as is reported in the main text) instead of the working age population (y-axis).

| **Table S11.** Sensitivity analysis of main effect from regression model (per 10% increase in population exposed to occupational heat) and estimated excess occupational heat deaths by exposure definition | | |
| --- | --- | --- |
| **Approach** | **Model coefficient** | **Excess deaths** |
| Among workers | 3.3% (1.0, 5.6) | 9,800 (3,100, 17,000) |
| Among working age population | 3.9% (0.9, 7.3) | 7,900 (1,700, 14,800) |

*No cross-county spillover effect.* In the first test, we scrambled estimated exposure prevalence across each county nationwide, but maintained the same temporal structure, 500 times and reran the quasi-Poisson model as specified in the main text (Eq. 2). In the second test, we restricted the scrambled exposure prevalences to within-metropolitan or nonmetropolitan areas, as defined by the US Bureau of Labor Statistics. For example, the Detroit-Warren-Dearborn metropolitan area consists of six counties (Wayne, Macomb, Oakland, Livingston, Lapeer, and St. Clair) due to shared economic activity around Detroit, Michigan. In the first test, the exposure prevalence of Wayne County in 2010 could be randomly assigned to any other county in the US in 2010 (e.g., Miami-Dade County, Florida); in the second test, it could only be randomly assigned to one of the six counties (i.e., Wayne, Macomb, … or St. Clair County) in 2010. In either test, if the occupational heat-mortality effects spillover to other counties, we would expect to observe an effect when estimating with shuffled exposure prevalence across counties. **Figure S11** suggests that any spillover effect is unlikely to be large, as we observe a null effect on average for each occupational risk factor when exposure prevalences are shuffled across counties in the same year, regardless of whether they were restricted (**Figure S11b**) or not (**Figure S11a**) to a metropolitan-nonmetropolitan area. This placebo test finds further evidence of consistency.

| **a** | 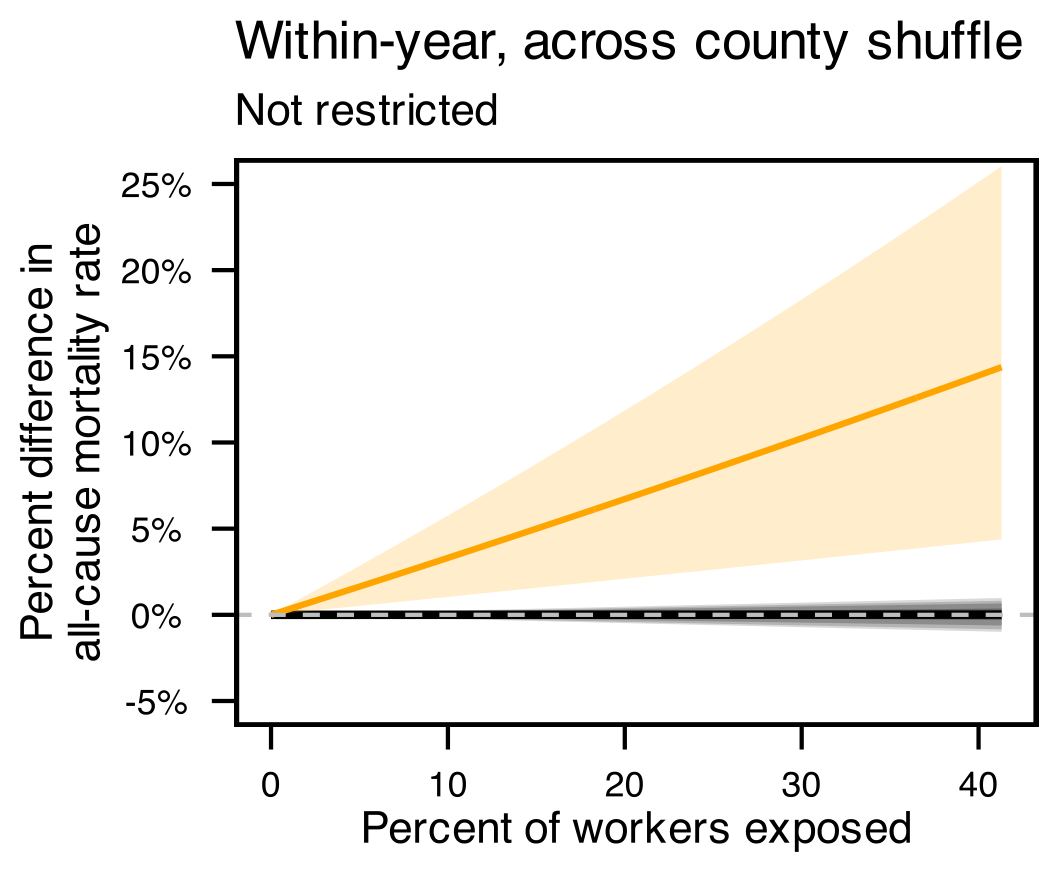 | **b** | 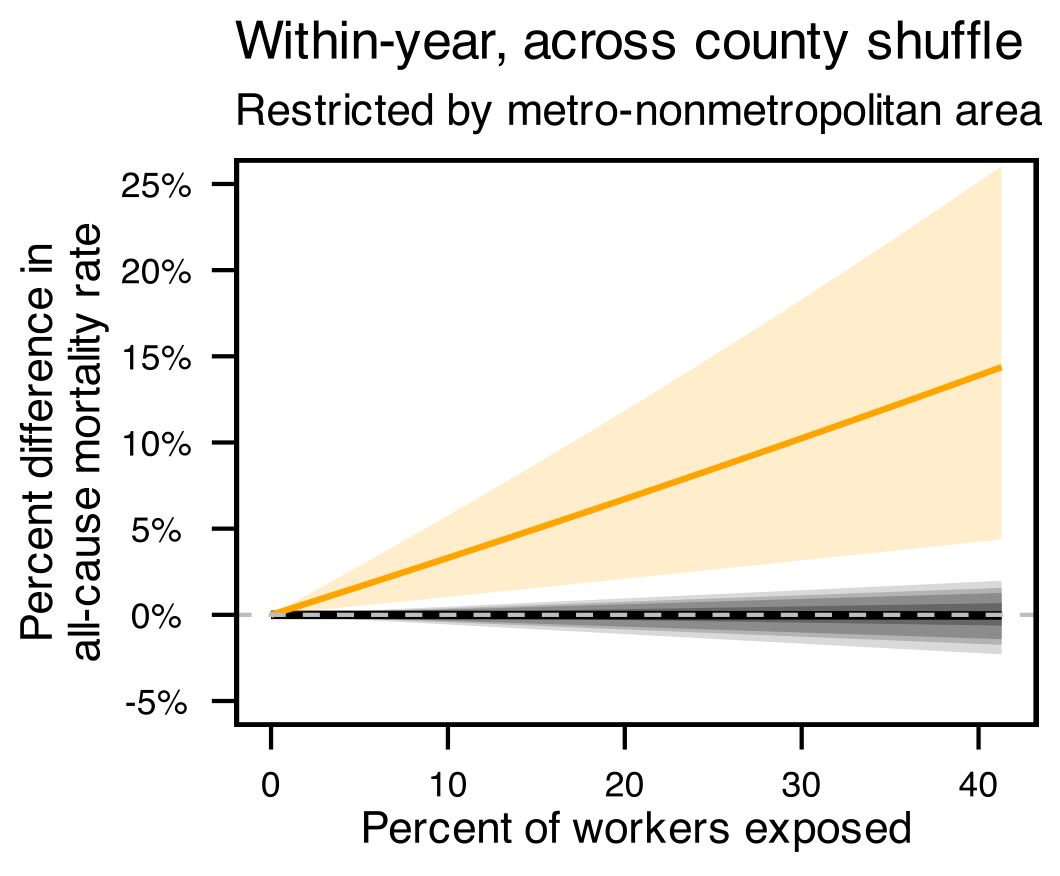 |
| --- | --- | --- | --- |

**Figure S11.** Within-year, across county randomization-based placebo test of the estimated occupational heat-mortality relationship. Left panel displays shuffling across all counties in the US, while right panel displays shuffling restricted to a county’s metropolitan-nonmetropolitan area. Shaded areas indicate the 2.5th, 5th, 10th, 25th, 75th, 90th, 95th, and 97.5th percentiles of the distribution across 500 randomized estimates using completely shuffled exposure prevalences, while the solid black line denotes the median of the randomized estimates. The orange line reproduces our central estimate (shaded orange area is 95% confidence interval) using the true, observed exposure prevalence data, as shown in **Fig. 3a**.
